# Distinct and shared genetic architectures of Gestational diabetes mellitus and Type 2 Diabetes Mellitus

**DOI:** 10.1101/2023.02.16.23286014

**Authors:** A. Elliott, R. K. Walters, M. Pirinen, M. Kurki, N. Junna, J. Goldstein, M.P. Reeve, H. Siirtola, S. Lemmelä, P. Turley, FinnGen, A. Palotie, M. Daly, E. Widén

**Author notes:** Corresponding authors Elisabeth Widén, Address: Institute for Molecular Medicine (FIMM), Biomedicum 2U, Tukholmankatu 8, 00290, Helsinki, Finland, Mark Daly, Address: Institute for Molecular Medicine (FIMM), Biomedicum 2U, Tukholmankatu 8, 00290, Helsinki, Finland.

## Abstract

Gestational diabetes mellitus (GDM) affects more than 16 million pregnancies annually worldwide and is related to an increased lifetime risk of Type 2 diabetes (T2D). The diseases are hypothesized to share a genetic predisposition, but there are few GWAS studies of GDM and none of them is sufficiently powered to assess whether any variants or biological pathways are specific to GDM. We conducted the largest genome-wide association study of GDM to date in 12,332 cases and 131,109 parous female controls in the FinnGen Study and identified 13 GDM-associated loci including 8 novel loci. Genetic features distinct from T2D were identified both at the locus and genomic scale. Our results suggest that the genetics of GDM risk falls into two distinct categories – one part conventional T2D polygenic risk and one part predominantly influencing mechanisms disrupted in pregnancy. Loci with GDM-predominant effects map to genes related to islet cells, central glucose homeostasis, steroidogenesis, and placental expression. These results pave the way for an improved biological understanding of GDM pathophysiology and its role in the development and course of T2D.

## Main text

Gestational diabetes mellitus (GDM) is a common heritable metabolic disorder characterized by new-onset glucose intolerance occurring during pregnancy and affecting 7-9% of women in the United States^1^ and 14% of pregnancies globally^2^. This, combined with adverse maternal and fetal outcomes in gestation, and subsequent high risk of maternal metabolic and cardiovascular disease with its associated mortality and morbidity, make GDM a major public health concern globally.

Clinically, GDM is related to an increased lifetime risk of Type 2 diabetes (T2D)^3-5^ – with over a third of women developing T2D within 15 years of their GDM diagnosis. GDM also aggregates in families and is associated with a family history of T2D^6-8^. Thus, it has been proposed that both diseases share a common etiology and a common genetic predisposition and that GDM would then merely represent a perturbation revealing an existing predisposition to T2D^9^. Yet, few studies have sought to uncover the genetic underpinnings of GDM and most of these studies have aimed at evaluating the impact of T2D loci^10-12^.

To date, only three Genome-Wide Association Studies (GWASes) have focused on the genetics of GDM^13-15^. The largest of them with 5,485 cases and 347,856 female controls revealed 5 genome-wide significant loci, of which 4 have also been previously associated with T2D^15^. While the results seem to support the hypothesis of shared etiology, none of the studies were sufficiently powered to directly assess the degree to which genetic risk is shared in GDM vs. T2D and whether any variants and biological pathways are specific to, or have differential effects in, GDM.

Thus, to elucidate the genetic underpinnings of GDM we conducted a genome-wide association study^16^ of GDM in 12,332 cases and 131,109 parous female controls. Participants were of Finnish ancestry from the FinnGen study. The association analysis was performed as described in Kurki et al^17^. Cases were identified using Finnish health and population registry sources including registry data from inpatient hospitalizations, outpatient specialty clinics, and birth registry. Cases were confirmed to have a diagnosis within a pregnancy window and those with diagnoses of diabetes prior to the index pregnancy were excluded (Online Methods, Supplementary Note).

Our GWAS nearly tripled the previously known loci for GDM, identifying 13 distinct associated chromosomal regions (Figure 1, Supplementary Figures 1-13). Fine mapping^18^ pinpointed 14 independent signals (the region near CDKN2A/B containing two independent signals) of which 9 regions had a 95% credible set containing 5 or fewer SNPs (Table 1, Supplementary Table 1, Online Methods). Nine regions represented novel GDM associations not reported in previous GDM GWAS. We further characterize the GDM GWAS results through annotation and colocalization of credible sets with >3,800 GWAS (Supplementary Tables 2-3), QTLs for gene expression, biomarkers and metabolites (Supplementary Tables 4-9), and chromatin interactions (Supplementary Table 10), along with tests of enrichment by functional consequence, gene expression, or canonical gene sets (Supplementary Tables 11-14; Supplementary Figures 14-17, Online Methods).

**Figure 1:**
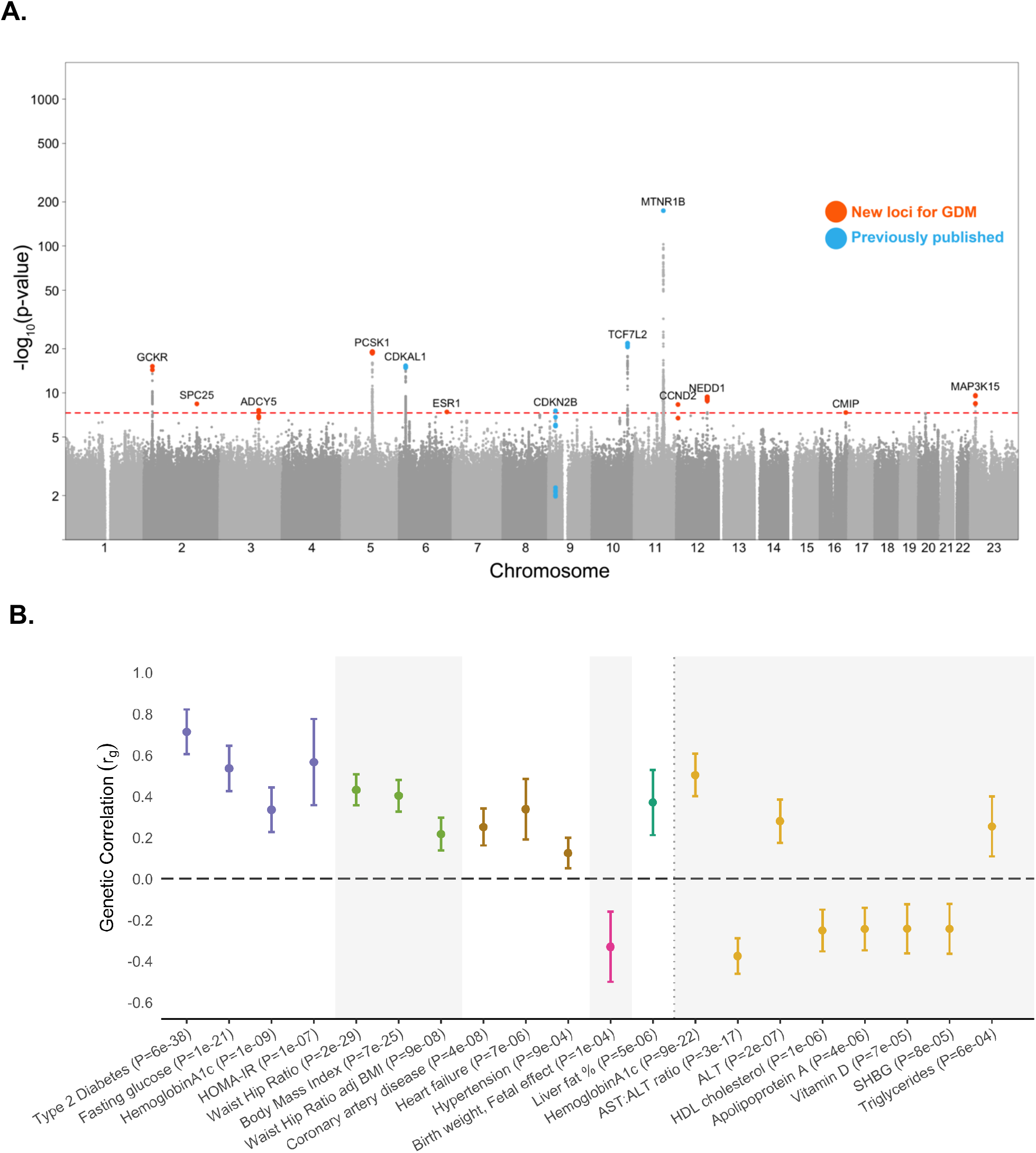
Genome-wide association results for GDM. (A) Manhattan plot of GWAS of GDM in 12,332 cases and 131,109 parous female controls of Finnish ancestry. The x-axis reflects chromosomal positions and the y-axis reflects −log10(P) values for the two-tailed association test for each variant, presented on a log scale. Red dotted line indicates the significance threshold (P = 5 × 10−8). Colored SNPs represent the credible set members for the 13 genome-wide significant loci, with blue indicating loci previously associated with GDM and orange indicating novel associations. Labels indicate the gene nearest to the fine-mapped lead SNP. (B) Genetic correlations (SNP-rg) between GDM and other diseases, traits and biomarkers estimated using LD score regression. Depicted traits are significant after Bonferroni correction for 53 traits (p < 9.4e-4); results for all tested traits are reported in Supplementary Tables 15-16. Error bars show +/- 1 standard error. Colors indicate phenotype category.

**Table 1:**
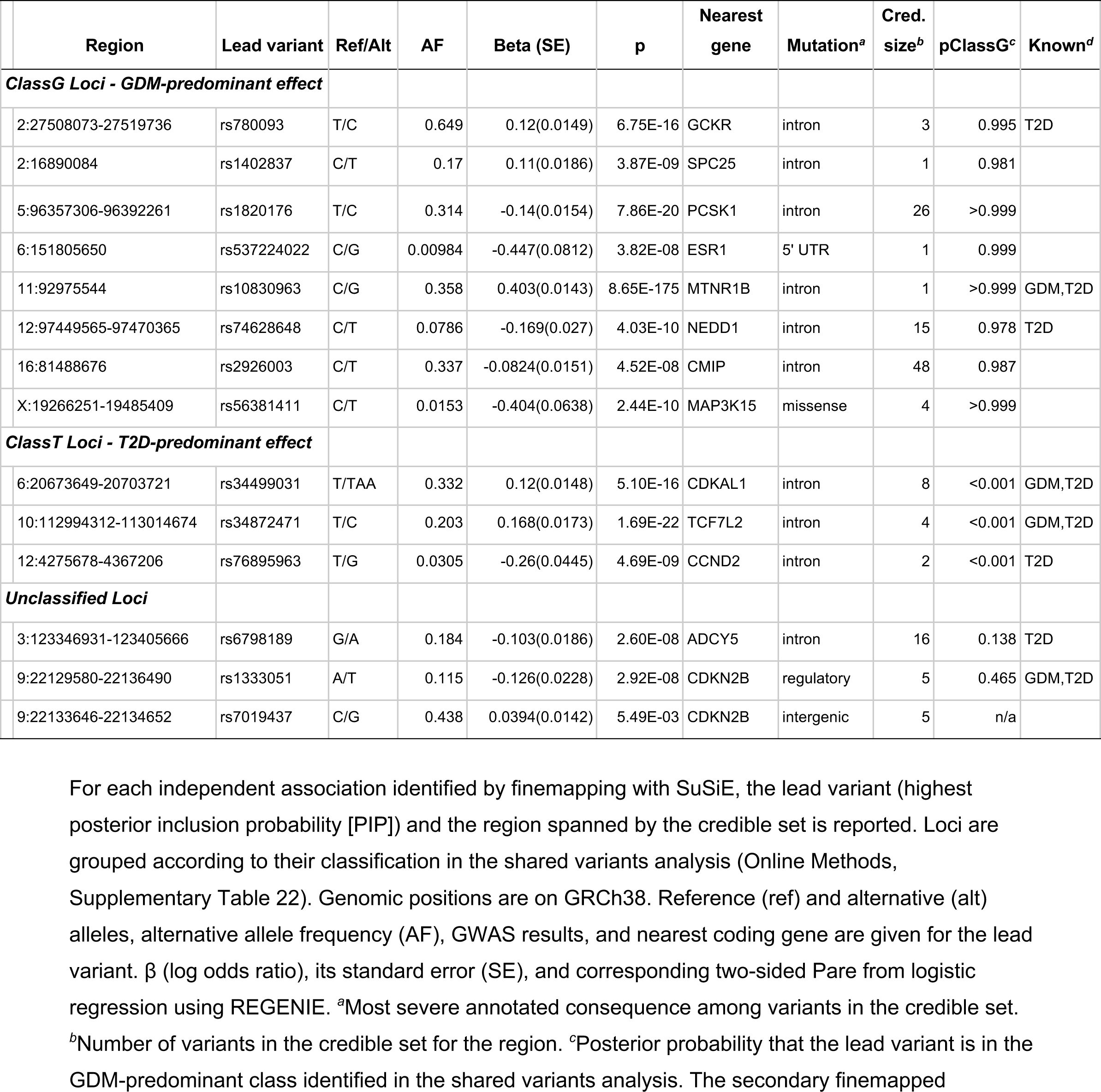
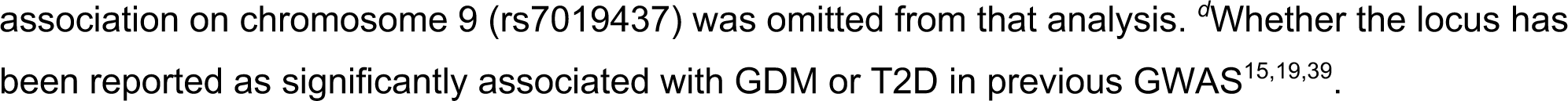
14 genome-wide significant fine-mapped signals for GDM.

| Region | Lead variant | Ref/Alt | AF | Beta (SE) | p | Nearest gene | Mutation<sup>a</sup> | Cred. size<sup>b</sup> | pClassG<sup>c</sup> | Known<sup>d</sup> |
| --- | --- | --- | --- | --- | --- | --- | --- | --- | --- | --- |
| ***ClassG Loci - GDM-predominant effect*** |  |  |  |  |  |  |  |  |  |  |
| 2:27508073-27519736 | rs780093 | T/C | 0.649 | 0.12(0.0149) | 6.75E-16 | GCKR | intron | 3 | 0.995 | T2D |
| 2:16890084 | rs1402837 | C/T | 0.17 | 0.11(0.0186) | 3.87E-09 | SPC25 | intron | 1 | 0.981 |  |
| 5:96357306-96392261 | rs1820176 | T/C | 0.314 | -0.14(0.0154) | 7.86E-20 | PCSK1 | intron | 26 | >0.999 |  |
| 6:151805650 | rs537224022 | C/G | 0.00984 | -0.447(0.0812) | 3.82E-08 | ESR1 | 5' UTR | 1 | 0.999 |  |
| 11:92975544 | rs10830963 | C/G | 0.358 | 0.403(0.0143) | 8.65E-175 | MTNR1B | intron | 1 | >0.999 | GDM,T2D |
| 12:97449565-97470365 | rs74628648 | C/T | 0.0786 | -0.169(0.027) | 4.03E-10 | NEDD1 | intron | 15 | 0.978 | T2D |
| 16:81488676 | rs2926003 | C/T | 0.337 | -0.0824(0.0151) | 4.52E-08 | CMIP | intron | 48 | 0.987 |  |
| X:19266251-19485409 | rs56381411 | C/T | 0.0153 | -0.404(0.0638) | 2.44E-10 | MAP3K15 | missense | 4 | >0.999 |  |
| ***ClassT Loci - T2D-predominant effect*** |  |  |  |  |  |  |  |  |  |  |
| 6:20673649-20703721 | rs34499031 | T/TAA | 0.332 | 0.12(0.0148) | 5.10E-16 | CDKAL1 | intron | 8 | <0.001 | GDM,T2D |
| 10:112994312-113014674 | rs34872471 | T/C | 0.203 | 0.168(0.0173) | 1.69E-22 | TCF7L2 | intron | 4 | <0.001 | GDM,T2D |
| 12:4275678-4367206 | rs76895963 | T/G | 0.0305 | -0.26(0.0445) | 4.69E-09 | CCND2 | intron | 2 | <0.001 | T2D |
| ***Unclassified Loci*** |  |  |  |  |  |  |  |  |  |  |
| 3:123346931-123405666 | rs6798189 | G/A | 0.184 | -0.103(0.0186) | 2.60E-08 | ADCY5 | intron | 16 | 0.138 | T2D |
| 9:22129580-22136490 | rs1333051 | A/T | 0.115 | -0.126(0.0228) | 2.92E-08 | CDKN2B | regulatory | 5 | 0.465 | GDM,T2D |
| 9:22133646-22134652 | rs7019437 | C/G | 0.438 | 0.0394(0.0142) | 5.49E-03 | CDKN2B | intergenic | 5 | n/a |  |

association on chromosome 9 (rs7019437) was omitted from that analysis. <sup>d</sup>Whether the locus has been reported as significantly associated with GDM or T2D in previous GWAS<sup>15,19,39</sup>.

We next performed analyses to evaluate the shared genetic etiology with T2D. Assessment of genome-wide significant signals using our algorithm Significant Cross-trait OUtliers and Trends in JOint York regression (SCOUTJOY; Online Methods) indicated that the 13 GDM-associated loci showed significant heterogeneity in their relationship to T2D (p< 0.001, Supplementary Table 21). Five of the 13 GDM-associated loci were not significantly associated (p<5e-8) with T2D in either a previously published large T2D meta-analysis^19^ or in FinnGen, while the remaining loci are established T2D hits (Table 1, Supplementary Figure 18). At the genomic level GDM and T2D were genetically correlated (rg=0.71, se= 0.06, p=6.8e-37), which is significantly greater than zero (p=6.8e-37) but less than 1 (p=1.2e-7; Online Methods). Significant genetic correlations were also seen with 12 diseases or traits and 8 blood laboratory values in cases where the disorder or value was phenotypically related to GDM (Figure 1; Supplementary Figures 19-20, Supplementary Tables 15-19). In both the genomic correlation and top hits comparison, GDM was significantly associated with fasting glucose, HbA1C, and 2 hour glucose result on oral glucose tolerance testing but was not associated with fasting insulin level. None of these glycemic traits or related disorders, however, appeared to stratify the 13 GDM-associated loci into distinct groups similar to T2D (Supplementary Figure 21, Supplementary Table 20).

To explore the relationship between GDM and T2D effects in more detail, we applied a Bayesian classification algorithm to the top associations for GDM and top associations for T2D (13 loci for GDM and 15 for T2D; Online Methods). The included T2D loci were the strongest signals obtained in a GWAS of male FinnGen participants (cases 27,607; controls 118,687) selected to have comparable statistical evidence for association (Supplementary Note). We utilized the male T2D scan to avoid confounding by sample overlap, though similar T2D results are seen in men and parous and nonparous women (Supplementary Note, Supplementary Tables 21 and 23-24, Supplementary Figure 22-24), suggesting the comparison is not confounded by overlap or sex-specific effects on T2D.

The shared variants analysis suggested that the genetics of GDM risk falls into two categories, one shared with T2D risk and the other uniquely gestational (Figure 2, Supplementary Table 22). Specifically, comparison of effect sizes between GDM and T2D does not support the existence of a single, consistent relationship between GDM and T2D across loci, but instead proposes two distinct classes of significant variants in this scan (Figure 2) – Class G, with GDM-predominant effects, and Class T, with T2D-predominant effects with the two-class model of relationship between GDM and T2D better fits the observed distribution of ORs by log10(likelihood-ratio) of approximately 30. Class G contains 8 of the 13 GDM-associated loci that have GDM-predominant SNP effects with an effect size roughly 3 times greater in GDM than T2D (Figure 2, Table 1). In comparison, the GDM-associated SNPs contained in Class T had effects in the two disorders that were consistent with T2D-signals significantly associated with diabetes only in the T2D GWAS – namely a reduced effect size in GDM versus T2D – a pattern of effects that was observed for all SNPs in Class T. The existence of the GDM-predominant class of effects, Class G, distinct from those traditionally seen in T2D, raises the possibility of physiologic mechanisms of glycemic control specifically important during pregnancy (Supplementary Note, Supplementary Table 25, Supplementary Figure 25).

**Figure 2:**
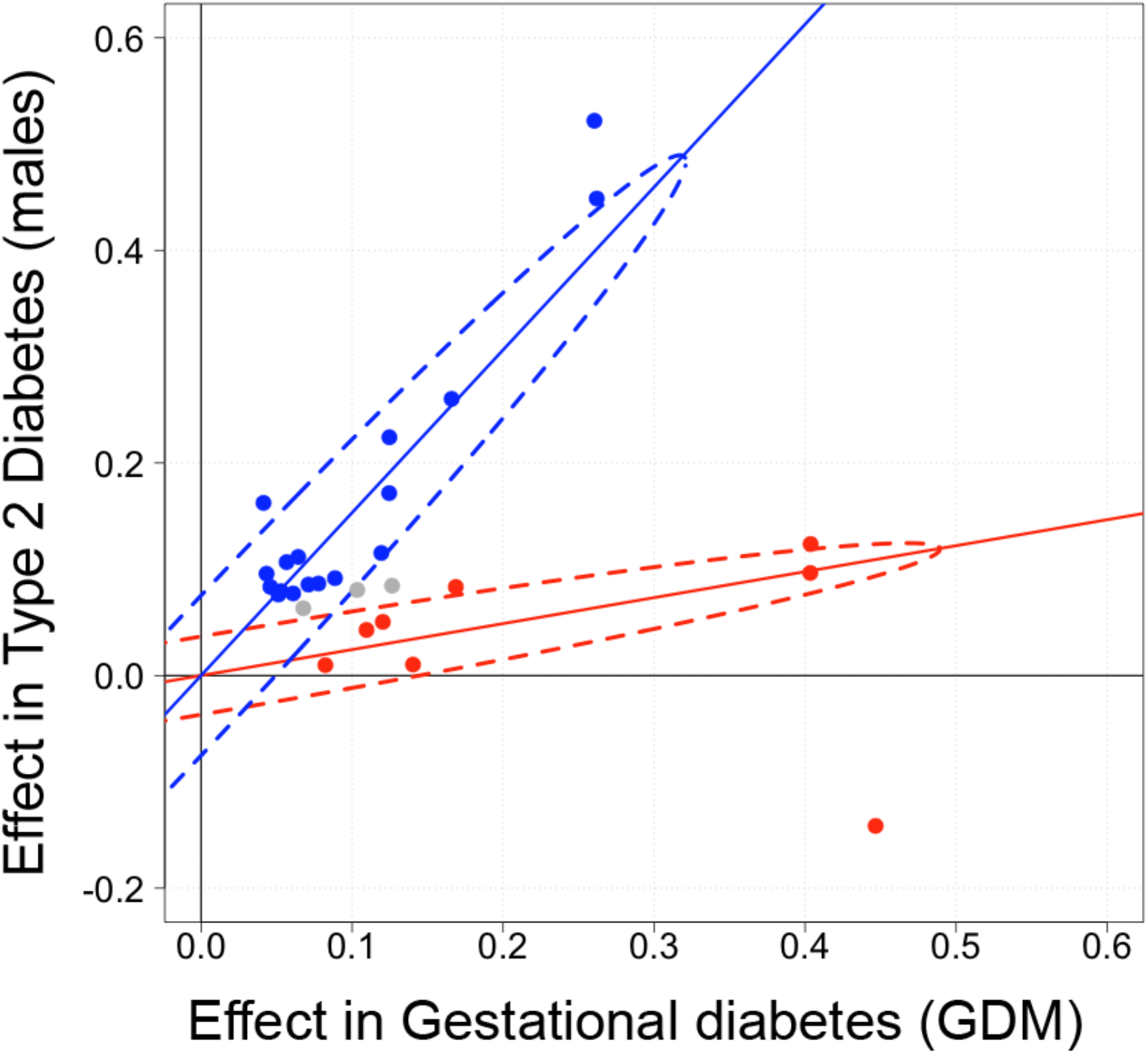
Classification of the genetic effects of SNPs in GDM and T2D. Comparison of log odds ratios in GWAS of GDM (x-axis) and T2D in males (y-axis) for top-associated SNPs from GDM (13 SNPs) and (15 SNPs). Two distinct classes of SNP effects were identified by a Bayesian classifier in shared variants analysis: Class T (blue) containing SNPs with T2D-predominant genetic effects and Class G (red) with GDM-predominant effects (Supplementary Table 22). Grey SNPs were not confidently assigned to either class (posterior probability > 95%). Dotted ellipses indicate the 95% probability regions of the fitted bivariate effect size distributions with each class.

As presented in Table 1, the eight class G loci have a peak SNP that is either intronic to a protein-coding gene, a missense mutation or, a 5’ UTR variant. Although the effects of a locus don’t always operate through the nearest gene, several of the loci implicate genes involved in plausible cellular processes e.g., signal transduction and hormone processing. (Table 1). For example, the missense mutation protective against GDM resides in *MAP3K15* on the X-chromosome (G838S, chrX_19380197_C_T_b38), which encodes a protein kinase (MAP3K15) that regulates apoptotic-mediated cell death and stress response. The gene has high expression in the adrenal glands and has previously been linked to steroidogenesis^20^ and polycystic ovarian syndrome^21^ The GDM-associated missense mutation is rare outside of Finland but other rare coding mutations in *MAP3K15* have previously been associated with T2D in UK Biobank^22^, i.e. female carriers of such rare nonsynonymous variants had a 30% reduced risk of T2D and reduced blood glucose and HbA1C levels, and hemizygous male carriers of rare protein-truncating variants had a 40% reduced risk of T2D. Further characterization of the GDM-associated mutation by PheWas analyses in FinnGen indicated that the mutation is associated with increased risk for hypertension (β = 0.11, p = 2.0e-8), but in contrast, there is no association with T2D (β = -0.09, p = 1.8e-3).

Other hormone-related class G signals include a variant mapping to the 5’UTR of the estrogen receptor gene, *ESR1*, and a variant nearby *PCSK1*, a gene encoding for prohormone convertase 1/3, which critically regulates endocrine and neuronal prohormone processing. Previous data show that homozygous loss of function of *PCSK1* results in a generalized and pleiotropic prohormone conversion defect characterized by severe obesity, impaired adrenal and thyroid function, reactive hypoglycemia, elevated levels of proinsulin, and low levels of insulin^23^, whereas common gene variants have been associated with BMI^24^, fasting proinsulin, fasting glucose, and T2D. Interestingly the GDM-associated allele identified in our study is associated with lower BMI (β = -0.02, p = 5.3e-11), lower weight (β = -0.02, p = 5.3e-11), and lower height (β = -0.01, p = 3.9e-6).

Finally, to gain further insight into potential functional differences between GDM and T2D we examined the cell type specific expression patterns associated with the GWAS summary statistics^25^ (Figure 3, Online Methods, Supplementary Tables 26-29, Supplementary Figures 26-28). We evaluate cell-type specific enrichment despite the lack of significant tissue-level enrichment because pregnancy induces major adaptive changes to specific cell populations within maternal tissues that might not be reflected in bulk tissue expression. Analyses integrating multiple large single cell RNA expression datasets indicated that pancreatic β cells are significantly associated both with GDM and T2D. However, only GDM had significant associations with the hypothalamus, i.e., hypothalamic GABAergic neurons (GABA2), hypothalamic glutaminergic neurons (GLU7), and neurons in the VMH arcuate nucleus (NR5a1_Adcyap1; Figure 3; Supplementary Table 28).

**Figure 3:**
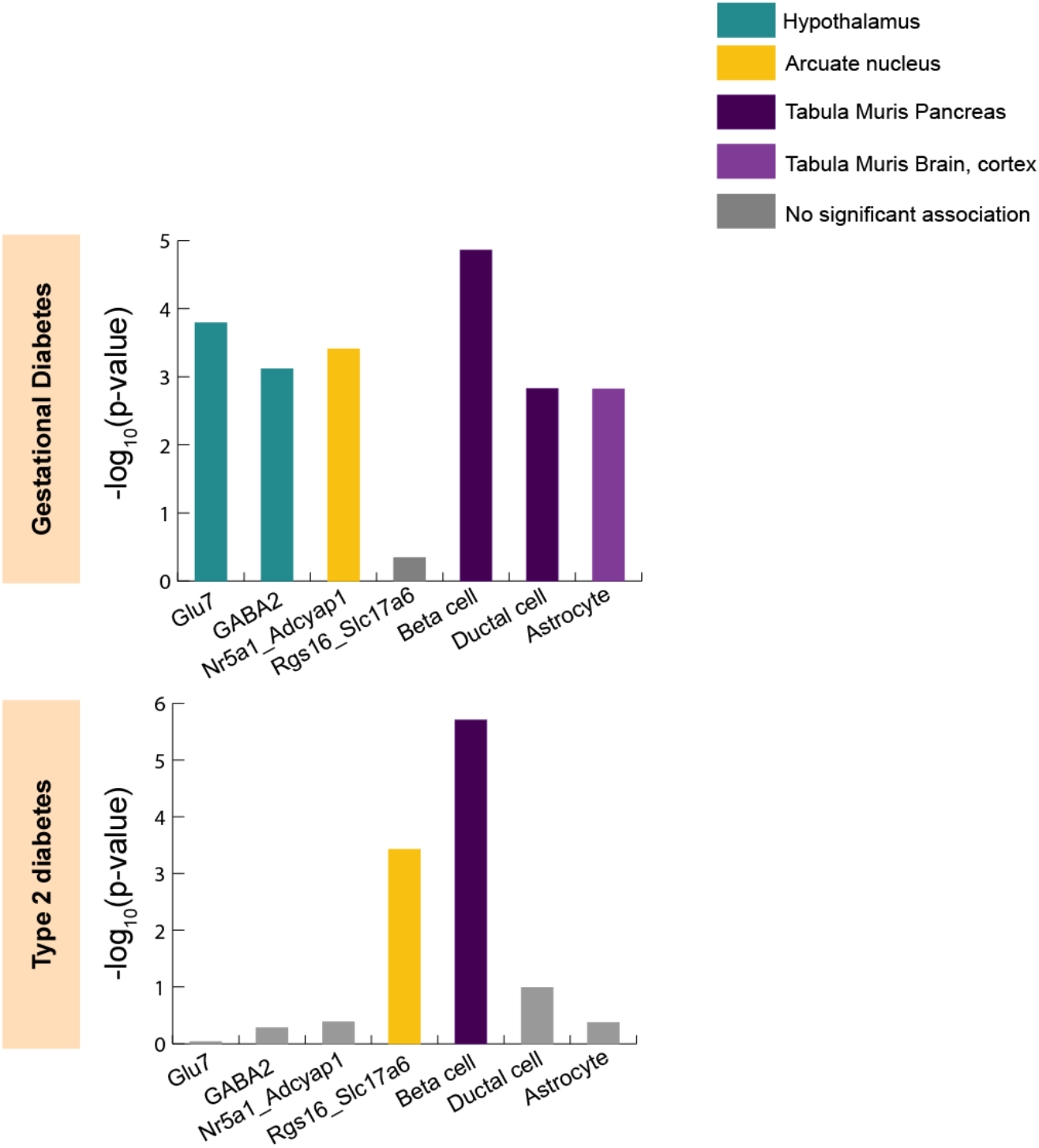
Cell type specificity analysis of GDM and T2D highlights different cell associations. Cell type specificity analysis was performed for GDM and for prior meta-analysis of T2D from Mahajan et al.^19^ using high quality murine single-cell RNA-seq datasets with FUMA (Supplementary Tables 26-29). Unadjusted P-values are reported for the association between relative gene expression in the given cell type and MAGMA gene-level associations in the GWAS. Results are shown for cell types that both (a) are significantly associated with at least one GWAS after correction for multiple testing of all cell types in all datasets, and (b) have putatively independent association conditional on other cell types in the same RNA-seq dataset. Colors indicate RNA-seq dataset source and significance.

Taken together, in this paper we present data from the largest GDM GWAS to date, identifying 14 independent signals in 13 associated chromosomal regions. GDM is a common disorder of pregnancy that has significantly increased in prevalence in all racial and ethnic groups in the last 15 years^26^. Despite conferring substantial morbidity to both mother and child, relatively little is known about the genetics of GDM outside of a proposed shared genetic etiology with T2D. Our key finding is that GDM in fact has a partially distinct genetic etiology, i.e., while GDM and T2D in part share a polygenic predisposition, there is a second category of GDM genetic risk factors that are predominantly gestational contributors to disease. This contextualizes the substantial effect of the *MTNR1B*, which had been reported previously as an outlier^11^, but our data now show that *MTNR1B* is representative of a whole group of GDM-predominant loci, characterized by a 3 times larger effect on GDM than on T2D.

Further studies will be required to characterize the precise GDM-specific molecular effects, but our current results suggest plausible mechanisms related to maternal adaptive physiological responses to pregnancy. Broadly, pregnancy increases circulating gestational hormones (e.g., human placental lactogen progesterone, and estrogen) causing alternations in normal homeostatic glycemic pathways in the brain and pancreas as well as impaired insulin sensitivity in maternal peripheral tissues. The brain and pancreas both show clear enrichment of signal in our cell-type specificity analysis of GDM, with our results in brain showing specific associations with hypothalamic and arcuate (ARC) neurons in GDM that are not seen in T2D (Figure 3, Supplementary Tables 26-28). Hypothalamic and ARC neurons have been implicated in adaptive glycemic response during pregnancy^27^. In that context, our ESR1 locus is particularly interesting given that the ventromedial hypothalamus (VMH) contains glucose-sensing neurons that express the estrogen receptor-α (ERα, encoded by ESR1) and act to regulate glucose levels^28^. Moreover, ERα neurons in the ARC have altered expression of several of our class G genes (e.g. *PCSK1, MTNR1B, SPC25*) in response to ERα knockout or perturbation of estrogen levels (which occurs in pregnancy)^29^. The involvement of these cells in GDM is further supported by our cell-type specificity results highlighting Nr5a1_Adcyap1 in ARC which project from the VMH^30^ and appear to correspond to a similar gene expression pattern to Glu7^31^ in our conditional analysis (Supplementary Figure 27, Supplementary Table 28, Supplementary Note). In contrast, while pregnancy is characterized by impaired insulin sensitivity and some of our class G genes (e.g. *PCSK1* and *MTNR1B*) affect insulin biosynthesis and secretion^32,33^, no evidence of a genetic relationship between GDM and insulin levels is observed either in top hits or genome-wide at the current sample sizes. Nonetheless, given the complexity of GDM the limited sample size of our dataset allows us to glean insight into only a few potential mechanisms. Much larger studies are warranted for a comprehensive view of the overall in-depth molecular underpinnings of GDM susceptibility.

The current study design in the rather homogeneous Finnish population carries specific strengths and weaknesses associated with this analysis approach. On one hand GWAS discovery is enhanced by population homogeneity^17^ and the linkage of national birth, inpatient and outpatient medical registries enables robust phenotyping (see Online Methods). The generalizability of the results may suffer however as some detected loci may be for rare alleles specifically enriched in the Finnish population. In our analyses of GDM two loci mapped to rare alleles enriched in Finland, which may be difficult to replicate elsewhere, while 70% of the loci correspond to variants that are common (MAF>10%), in non-Finnish European ancestry individuals (Table 1). Nonetheless, additional studies prioritizing ancestrally diverse populations are needed for a better understanding of the genetic underpinnings of GDM in all populations at risk.

In summary, we discovered 9 novel loci associated with GDM and demonstrate that GDM genetic risk is distinct from T2D both at the locus and genomic scale. Our results suggest that the genetics of GDM risk falls into two categories – one part T2D risk and one part predominantly gestational contributors to disease. Tissue characterization of GDM genetics further implicates tissues previously identified in adaptive pregnancy responses, raising hypotheses regarding genetic effects in these tissues during pregnancy. Broadly this work underscores the benefits of focusing on resources on disorders of pregnancy as pregnancy is a natural perturbation that offers leverage to discover loci with novel physiologic mechanisms of glycemic or homeostatic control.

## Methods

### Cohort

The FinnGen Study is a public–private partnership project combining data from Finnish biobanks and electronic health records from national registries. The linked national health registers include data on hospital and outpatient visits, primary care, cause of death, and medication records. Approval from the FinnGen Study was received to use the data in the present work. After a 1-year embargo, the FinnGen summary stats are available for download. In this study, we used the results from the FinnGen release R8, which includes data from 342,499 individuals and more than 4500 disease endpoints.

### Phenotyping

Full details of phenotyping are described in the Supplementary Note. Briefly, clinical endpoints with corresponding dates were constructed for gestational diabetes and related diagnoses for exclusions for all FinnGen participants as described in the Supplementary Note. Temporal phenotyping was then performed to phenotype each pregnancy for presence of glycemic disease and then assign individuals as cases or controls. Beginning with 330,000 pregnancies among genotyped FinnGen participants, we defined a “pregnancy window” of 40 before delivery until 5 weeks after delivery. A pregnancy met inclusion criteria for “gestational diabetes” if it had (I1) gestational diabetes ICD codes occurring in the pregnancy window, (I2) any diabetes codes occurring in the pregnancy window (e.g. for ICD8), or (I3) abnormal blood glucose test results in the Medical birth register, which contains data on the mother’s diseases during pregnancy. Pregnancies were then excluded for: (E1) any previous diabetes diagnosis code occurring outside a pregnancy window; (E2) any previous significant pancreatic disease, including chronic pancreatitis, pancreatic necrosis, pancreatic cancer, or cystic fibrosis; or (E3) any previous Type 1 or Type 2 diabetes code. Pregnancies passing these exclusion criteria and without any inclusion criteria for gestational diabetes were designated as “unaffected”. Then to phenotype individuals, cases were identified among the 151,000 genotyped females with a history of pregnancy as those at least 1 pregnancy meeting inclusion criteria for gestational diabetes and passing exclusion criteria. Controls were defined as females with only “unaffected” pregnancies (i.e. no diabetes or significant pancreatic diseases occurring prior to or during any pregnancy, and no abnormal blood glucose in the Medical birth register).

### Genotyping and GWAS

A detailed description of the study design and analytical methods are available in the online documentation.

In brief, FinnGen individuals have been genotyped with Illumina and Affymetrix chip arrays. QC was performed to remove samples and variants of poor quality. Imputation was performed using population-specific SISu v3 importation reference panel.

A subset of unrelated individuals of genetically confirmed Finnish ancestry was identified. GWAS was performed using REGENIE 2.2.4. Sex, age, 10 PCs, and genotyping batch were included as covariates in the analysis.

### Finemapping

Finemapping of a 1.5MB locus around any GWAS lead SNP was performed using the SuSiE algorithm^18^ which reports causal variants and a 95% credible set for each independent signal. Details described previously^17^ and here, https://finngen.gitbook.io/documentation/methods/finemapping. As LD, we used in-sample dosages (i.e cases and controls used for each phenotype) computed with LDStore. Independent signals were those that either represent the primary strongest signal with lead p< 5*10-8 or as secondary signals that must have genome-wide significance and log bayes factor (BF)>2.

### Annotation

Variants were annotated with Ensembl Variant Effect Predictor (VEP) version 104, (https://www.ensembl.org/info/docs/tools/vep/index.html) data to give the projected variant consequence. Each variant was also annotated for enrichment in Finland compared to compared to non-Finnish-Swedish-Estonian Europeans, as described previously^17^. Annotation with known prior GWAS loci was performed as previously described^17^. In brief, for each independent association we annotated every phenotype in GWAS Catalog that was significantly associated with either (1) the lead PIP variant or (2) any variant in the credible set. Similar annotation was performed for metabolite associations from the MetSIM study^34^ (Supplementary Note). Each locus was also annotated with SNP2GENE in FUMA version v1.3.7 (fuma.ctglab.nl/snp2gene/) for chromatin interactions (Supplementary Table 10), eQTL associations (Supplementary Table 4), and prior GWAS hits (Supplementary Table 3).

### Colocalization

Colocalization was performed on all fine mapped regions as previously described for the FinnGen study^17^. In brief, the probabilistic model to intergrate GWAS and eQTL data was eCAVIAR^35^ however the input posterior inclusion probabilities (PIP) estimated by the SuSiE algorithm^18^. The eCAVIAR method uses PIPs for variants in each region to compute a colocalization posterior probability (CLPP, see Supplementary Note). The intersection of variants in credible sets was then checked across multiple phenotypes from FinnGen (Supplementary Table 2), GTEx (Supplementary Table 5), eQTL Catalogue (Supplementary Table 6), GeneRisk (Supplementary Table 7), and the UK Biobank (Supplementary Table 8).

### Gene enrichment analysis

Gene-level association results from MAGMA were used to identify tissue and pathway enrichments using the SNP2GENE and GENE2FUNC modules of FUMA (version v1.4.1). The MAGMA results were tested for (a) association with gene expression levels in GTEx v8 (Supplementary Table 12, Supplementary Figure 14), (b) enrichment in sets of differentially expressed genes identified across tissues from GTEx v8 (Supplementary Table 13, Supplementary Figure 15), (c) enrichment in gene sets for pathways or other biological processes including those defined by KEGG (MsigDB c2), GO biological processes (MsigDB c5), or WikiPathways (Supplementary Table 14), and (d) enrichment in gene sets defined by reported associations in GWAS Catalog (Supplementary Table 14, Supplementary Figure 17).

### Genetic correlation

We estimated the SNP heritability (*h*^2^_SNP_) of GDM and pairwise genetic correlations (SNP-r_g_) between GDM and diabetes-related diseases and traits using LDSC Version 1.0.1. Testing difference of *r*_*g*_ from perfect correlation was performed using a one tailed Z-score test:

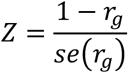

See Supplementary Note for details on additional genetic correlation analyses.

### Significant Cross-trait OUtliers and Trends in JOint York regression (SCOUTJOY)

To compare heterogeneity of GDM-associated loci’s genetic effects in any two disorders we developed SCOUTJOY (Supplementary Note), extending the heterogeneity detection algorithm introduced by MR-PRESSO^36^ to handle sample overlap and estimation error in GWAS of each phenotype. To estimate the primary relationship in effect sizes between the two disorders while accounting for estimation error we derive estimators for York regression^37^ with a fixed intercept. Global heterogeneity testing was then performed using simulated null replicates to test the hypothesis that the observed effect sizes across top hits are consistent with a single uniform relationship. Outlier variants were similarly identified as having larger than expected residuals from the York regression. We extend this outlier detection method from MR-PRESSO to allow iterative assessment of each variant based on the outlier status of all other variants. Code for SCOUTJOY and York regression with a fixed intercept is available on github (link to be provided on acceptance).

### Shared variants analysis

We applied linemodels package (https://github.com/mjpirinen/linemodels) to summary statistics from T2D GWAS and GDM GWAS. The analysis included 28 lead variants from the GWASes (13 from GDM and 15 from T2D). We classified the variants into two classes based on their bivariate effect sizes. The classes were represented by line models whose slopes were estimated using an EM-algorithm, resulting in values 1.53 (labelled as class T) and 0.25 (labelled as class G). For both models, the scale parameters determining the magnitude of effect sizes were set to 0.2 and the correlation parameters determining the allowed deviation from the lines were set to 0.99. The membership probabilities in the two classes were computed separately for each variant by assuming that the classes were equally probable a priori. Since the two GWAS did not have overlapping samples, the correlation of their effect estimators was set to 0.

### Cell type specificity analyses

To get better resolution on specific cell types, we performed cell-type specificity analyses with high quality single cell murine datasets using FUMA (https://fuma.ctglab.nl/tutorial#celltype; Supplementary Note). First, we identified tissue-level associations with Tabula Muris data^38^ identifying significant associations (FDR < .05) with expression in brain and pancreas after Benjamini-Hochberg multiple testing correction (Supplementary Figure 27). We then performed cell-type specificity analyses as previously described^25^, augmenting Tabula Muris with additional high quality scRNA-seq of hypothesized involved brain regions (Supplementary Note). Analysis was performed on genetic summary statistics for both our Gestational Diabetes GWAS and for a recent Type 2 Diabetes European meta-analysis dataset^19^. We also compare the pancreatic results to analysis of high quality scRNA-seq of pancreas in humans to assess the impact of known differences in human vs. mouse pancreatic cellular function and physiology (Supplementary Table 29, Supplementary Figure 28).

### Ethics statement

Patients and control subjects in FinnGen provided informed consent for biobank research, based on the Finnish Biobank Act. Alternatively, separate research cohorts, collected prior the Finnish Biobank Act came into effect (in September 2013) and start of FinnGen (August 2017), were collected based on study-specific consents and later transferred to the Finnish biobanks after approval by Fimea, the National Supervisory Authority for Welfare and Health. Recruitment protocols followed the biobank protocols approved by Fimea. The Coordinating Ethics Committee of the Hospital District of Helsinki and Uusimaa (HUS) approved the FinnGen study protocol Nr HUS/990/2017.

The FinnGen study is approved by Finnish Institute for Health and Welfare (THL), approval number THL/2031/6.02.00/2017, amendments THL/1101/5.05.00/2017, THL/341/6.02.00/2018, THL/2222/6.02.00/2018, THL/283/6.02.00/2019, THL/1721/5.05.00/2019, Digital and population data service agency VRK43431/2017-3, VRK/6909/2018-3, VRK/4415/2019-3 the Social Insurance Institution (KELA) KELA 58/522/2017, KELA 131/522/2018, KELA 70/522/2019, KELA 98/522/2019, and Statistics Finland TK-53-1041-17.

## Data Availability

FinnGen summary data (genome-wide association summary statistics and phenotype data) are made freely available every 6 months. The FinnGen data may be accessed through Finnish Biobanks' FinnBB portal (www.finbb.fi) and THL Biobank data through THL Biobank (https://thl.fi/en/web/thl-biobank).

## Availability Statement

FinnGen summary data (genome-wide association summary statistics and phenotype data) are made freely available every 6 month. The FinnGen data may be accessed through Finnish Biobanks’ FinnBB portal (www.finbb.fi) and THL Biobank data through THL Biobank (https://thl.fi/en/web/thl-biobank).

