## Supplemental text for "Distinct and shared genetic architectures of Gestational diabetes mellitus and Type 2 Diabetes Mellitus"

### Supplementary Text

#### I. Phenotyping

Registry data is available for all FinnGen participants from national health registers. This includes data on hospital and outpatient visits (HILMO - Care Register for Health Care), Primary care visits (AvoHILMO - Register of Primary Health Care), the Medical Birth Register (including data on the mother's diseases during pregnancy: ICD-9,10), Causes of Death, reimbursed medication entitlements and prescribed medicine purchases, and the Finnish Cancer Registry. Clinical endpoints were constructed from the register codes using Finnish version of International Classification of Diseases, 10th revision (ICD-10) diagnosis codes and harmonizing those with definitions from ICD-8 and ICD-9.

Codes related to diagnoses are as follows:

| Phenotype | Codes |
|---|---|
| Gestational diabetes | *ICD10*: O244;<br><br>*ICD9*: 6480A;<br><br>*ICD8*: None |
| Diabetes | *ICD10*: E10, E10.[0-9], E11, E11.[0-9], E13, E13.[0-9], E14, E14.[0-9], M14.2, H36.0[0-9], O24, O24.[1-4];<br><br>*ICD9*: 250, 250.[0-8]A, 250.[0-8]B, 250.[0-8]C, 250.[0-8]X, 3620A, 3620B, 6480A, 6488A;<br><br>*ICD8*: 250, 2500[0-9],76110;<br><br>KELA drug reimbursement registry: 103, 215;<br><br>Drug ATC code: A10AB0[1-6], A10AB30, A10AC0[1-4], A10AC30, A10AD0[1-6], A10AD30, A10AE0[1-6], A10AE30, A10AE54, A10AE56, A10BA, A10BA0[1-3], A10BB, A10BB0[1-9], A10BB1[0-2], A10BB31, A10BC, A10BC01; |
| Chronic pancreatitis and pancreatic necrosis | *ICD10*: K86.01, K86.08, K86.1;<br><br>*ICD9*: 5771A, 5771B, 5771C, 5771D;<br><br>*ICD8*: 57710, 57719, 57702,57793; |
| Pancreatic cancer | *ICD10*: C25, C25.[0-9];<br><br>*ICD9*: 157, 157[0-4]A, 157[8-9]X;<br><br>*ICD8*: 157, 15701, 15787, 15788, 15799, 23060; |
| Cystic fibrosis | *ICD10*: E84, E84.[0-9];<br><br>*ICD9*: 2770A;<br><br>*ICD8*: 27300; |

Temporal phenotyping was then performed in 2 steps: (1) pregnancies with gestational diabetes were identified, (2) individual females were assigned as cases or controls.

Date of delivery/ the child's birth date of 330,000 children born to female FinnGen participants was identified from the Population Register or the Medical Birth Register. Then a "pregnancy window" was generated for each pregnancy from 40 weeks before delivery to 5 weeks after. The allotted timing was to account for variability introduced by pseudoanonymization of time scale within and across registries. A pregnancy was classified as having "gestational diabetes" or "unaffected" based on the following criteria:

| | Inclusion Criteria | Exclusion Criteria |
|---|---|---|
| Pregnancy with GDM | Pregnancies were included if they had any of the following:<br><br>1. Abnormal blood glucose listed in the pregnancy registry<br>2. Gestational diabetes code[†] received in the pregnancy window<br>3. Diabetes code received in the pregnancy window | Pregnancies were excluded if any of the following occurred prior to the index pregnancy:<br><br>1. Significant pancreatic disease including:<br>• Chronic pancreatitis<br>• Pancreatic necrosis<br>• Pancreatic cancer<br>• Cystic fibrosis<br>2. Diabetes diagnosis code occurring outside a pregnancy window<br>3. Type 1 or Type 2 diabetes[†] at any point prior |
| Unaffected pregnancy | None of the GDM inclusion criteria | Same as above |
| [†]Recall these definitions contain only ICD9 and ICD10 codes. | | |

The inclusion criteria reflect that a gestational diabetes diagnosis code was only available in ICD9 and ICD10 whereas in ICD8 there was only a general "diabetes" code. To include representation from ICD8 era, we constructed the inclusion and exclusion criteria to pull females who received a diagnosis of diabetes during a pregnancy window who had no other diabetes codes prior to pregnancy. The vast majority of cases were from ICD9 and ICD10 with only 730 cases added. Comparison of pregnancy labels created with the addition of ICD8 to the cohort to those without demonstrated they were sufficiently similar to proceed.

Among the 151,000 genotyped females with a pregnancy, cases were identified as those with gestational diabetes in at least 1 pregnancy. Controls are females with a history of pregnancy where all pregnancies were "unaffected". In other words, controls have (1) no abnormal blood glucose in the pregnancy registry, (2) no diabetes codes occurring prior to or during any pregnancy, and (3) no significant pancreatic disease prior to or during any pregnancy.

#### II. Colocalization comparison metrics

We perform colocalization on finemapped credible sets obtained from SuSiE using the posterior inclusion probabilities (PIP) estimated by SuSIE. For the colocalization we report both the Causal Posterior Probability (CLPP) and the Causal Posterior Agreement (CLPA). Colocalization was performed for the FinnGen release 8 endpoints (Supplementary Table 2), gene expression data from GTEx (Supplementary Table 5) and the eQTL Catalogue (Supplementary Table 6), lipid measures (Supplementary Table 7) and biomarkers (Supplementary Table 8).

##### A. Causal Posterior Probability (CLPP)

The Causal Posterior Probability (CLPP) is computed similarly to Hormozdiari et al (equation 8)[1]. For the credible set of phenotype 1 ($cs_1$) and 2 ($cs_2$).

$$CLPP = \sum_{i \in cs_1 \cap cs_2}^{i} x_i * y_i$$

where x and y are vectors containing the PIP for $cs_1$ and $cs_2$, respectively. Notably CLPP is dependent on the credible set size.

##### B. Causal Posterior Agreement (CLPA)

The Causal Posterior Agreement (CLPA) is independent of credible set size and defined by

$$CLPA = \sum_{i \in cs_1 \cap cs_2}^{i} \min\left(x_1 * y_i\right)$$

##### C. Datasets

Colocalization was performed for SuSiE fine-mapped data from: FinnGen phenotype endpoints from FinnGen release 8 (Supplementary Table 2); gene expression data from 49 tissues from donors of mixed ancestry in GTEx v8 (Supplementary Table 5)[2]. Gene expression data from EMBL-EBI eQTL Catalogue on 24 tissues/cell types[3] from samples of predominantly European ancestry (Supplementary Table 6); metabolism eQTL datasets from the GeneRISK study[4,5], including 186 lipid species (Supplementary Table 7); and 36 continuous endpoints and 57 biomarkers from UK Biobank(Supplementary Table 8);.

#### III. Finnish metabolite mQTL annotations

We annotate the SuSiE finemapped credible sets for GDM and Type 2 diabetes with metabolite QTLs (mQTL) on circulating metabolite data from the Finnish METSIM study[6] .

##### A. Dataset

The Metabolic Syndrome in Men (METSIM) study[6] is a Finnish population study 10,197 Finnish men examined in 2005-2010. Genetic data includes both array data and exome data. Fasting blood metabolites were obtained with NMR mass spectrometry. Notably, this is an all male study which is a potential limitation for our GDM annotations however this is in the same population as our study. Every SNP in METSIM has been tested for association with 1391 metabolites, 937 after removing metabolites that are classified as xenobiotics or uncharacterized.

##### B. Method

We evaluated the 28 diabetes loci (the 13 GDM associated loci and 15 loci from the T2D GWAS used in the shared variants analysis) to annotate the SuSiE finemapped credible sets with mQTLs on circulating metabolite data from the METSIM study. To annotate the loci, we choose a representative lead SNP for the metabolite annotation (metLead variant) that is the credible set variant with the highest SuSiE posterior inclusion probability, PIP, that occurs in the METSIM study. If no credible set member appears in METSIM then we assign the metLead variant as the variant in METSIM with the highest LD to the lead variant if the r2 is > 0.2, otherwise assign NA.Associations with 937 blood metabolites were then extracted for each of the metLead SNPs. Metabolites with significant p-values after Bonferroni correction were then annotated to the locus (threshold 5.3e-5 for multiple testing of 937 metabolites).

##### C. Results

We provide mQTL annotations for all of the GDM loci and for T2D loci. Seven of the 13 GDM loci have mQTL annotations. Novel GDM loci are associated with glucose, mannose, branched chain amino acids, serine, androgens, bile acids, and lipids. Detailed annotations provided in Supplementary Table 9.

#### IV. Comparison of genetic correlation results

Gestational diabetes was tested for genetic correlation ($r_g$) with 24 disorders or traits and with 29 biomarkers. Significantly correlated traits or biomarkers had their $r_g$ in GDM compared to the $r_g$ with T2D to determine if there was a significantly different overlap with T2D than GDM.

##### A. Method to compare genetic correlation results

For both GDM and T2D, initial pairwise genetic correlations (SNP-$r_g$) between GDM and other traits was performed using LD Score Regression (LDSC) Version 1.0.1 with previously computed LD scores from European ancestry individuals from the 1000 Genomes Project. We then compared the magnitude of SNP-$r_g$ that was seen for each trait with GDM vs T2D by computing the Z-score for the comparison in SNP-$r_g$ between trait 1 and 2 as follows:

$$Z = \frac{r_{g,gdm} - r_{g,t2d}}{\sqrt[2]{se\left(r_{g,gdm}\right)^2 + se\left(r_{g,t2d}\right)^2}}$$

P-values for the comparison were computed from the Z-score.

##### B. Results

Significant genetic correlations were identified with 12 diseases or traits and 8 blood laboratory values all of which had been phenotypically related to GDM (Supplementary Table 15 & 16; Figure 1b). The 12 traits with significant genetic correlation included glycemic traits such as fasting glucose, Hb1C, HOMA-IR, and 2 hour glucose on OGTT testing but not Fasting insulin (Supplementary Table 15). Adulthood disorders in which we observed significant rg to GDM like Coronary Artery Disease (CAD), Heart Failure (HF), and Hypertension (HTN) have previously been phenotypically linked to gestational diabetes[7-9]. Significant biomarkers included liver and lipid biomarkers linked to GDM[10-12]as well as apolipoprotein A and Vitamin D that have recently been implicated prior to or early in GDM pregnancies[13-15].

We then took the eleven traits with significant genetic correlation (excluding T2D) and compared the $r_g$ in GDM to the $r_g$ in T2D, (Supplementary Table 17 and Supplementary Figure 19). Three traits had significantly different $r_g$ with GDM than with T2D after multiple testing correction: Waist Hip Ratio, BMI, and CAD. Comparison of the $r_g$ to GDM versus to T2D in 20 biomarkers showed a significant difference in the genetic correlation of two biomarkers after correction for multiple testing: urea and gamma glutamyltransferase (Supplementary Table 18 and Supplementary Figure 20).

In particular, maternal GDM is known to affect child birth weight. Genetic studies of birth weight have identified both fetal and maternal genetic associations with the child birth weight[16] and have used structural equation modelling to distinguish the separate contributions of maternal and fetal effects. Based on the observed genetic correlation of these maternal and fetal effects with fasting glucose it was hypothesized that genetic signals for GDM in the mother would be positively corelated with child birth weight while genetic signals for GDM in the fetus would be negatively correlated with fetal birth weight. Intuitively this is because glucose but not insulin passes the placenta and fetal insulin is an important growth hormone stimulating fetal size and development. Thus a fetal genetic effect that results in reduced GDM risk was thought to also correspond to reduced insulin levels and reduced production of this hormone essential for growth. Our GDM GWAS confirms this hypothesis that GDM has a significant positive genetic association with maternal GWAS of birthweight and maternal effect on birth weight but a negative genetic correlation with fetal GWAS of birth weight and fetal effect on birth weight (Supplementary Table 19, Supplementary Figure 19). However contrary to the hypothesis in the paper, we do not see any significant association to fasting insulin levels at current sample sizes, suggesting any genetic correlation of GDM with general fasting insulin levels is likely modest (Supplementary Table 15&20).

### V. Comparison of GDM with T2D and glycemic traits

Initial evaluation of the top SNPs for GDM comparing their genetic effects in GDM and T2D suggested a relationship as in Supplementary Figure 18. To more formally compare the genetic effect of 13 GDM-associated loci on GDM versus T2D and 6 other disorders, we extended existing methods[17-20] to determine trends of effects across the loci and identify outliers that have different pattern of effects than the majority of SNPs. Our approach, Significant Cross-trait OUtliers and Trends in JOint York regression (SCOUTJOY), has the advantage of: (1) accounting for sampling error in the GWAS effect estimates for each phenotype, (2) allowing sample overlap or other sources of correlated sampling error, and (3) making no assumptions about a causal relationship between the phenotypes. To accomplish this we first had to derive estimating equations for a special case of York regression that has not been addressed by existing literature. Using this tool we were able to compare the loci's genetic effects in GDM vs glycemic traits including T2D and the glycemic traits such as fasting glucose, 2h glucose on Oral Glucose Tolerance Testing (OGTT), and fasting insulin.

| Variables | |
|---|---|
| $X_i$, $Y_i$ | GWAS effect sizes for SNP *i* for the two diseases/traits being compared |
| i | Index for chosen top SNPs 1,…,$K$ |
| K | Total number of SNPs being compared |
| $\sigma_{x,i}$ , $\sigma_{y,i}$ | Standard errors of $X_i$ and $Y_i$ for SNP *i* |
| $\rho_i$ | correlation of the errors for $x_i$ and $y_i$ |
| a | Intercept of regression line comparing effect sizes between two traits |
| $a_0$ | Chosen fixed intercept for regression |
| b | Slope of the regression line comparing effect sizes between two traits |
| $W_i$ | Weights $$\frac{1}{\sigma_{y,i}^2 + b^2\sigma_{x,i}^2 - 2b\rho_i\sigma_{x,i}\sigma_{y,i}}$$ |
| $\bar{X}, \bar{Y}$ | Weighted means of X and Y $$\overline{X} = \frac{\sum_{i=1}^{k} W_i\,\widehat{X_i}}{\sum_{i=1}^{k} W_i}, \overline{Y} = \frac{\sum_{i=1}^{k} W_i\,\widehat{Y_i}}{\sum_{i=1}^{k} W_i}$$ |
| $\widehat{X_i}, \widehat{Y_i}$ | Estimate of the effect size under the York regression slope |
| M | Number of simulations |
| $\alpha_{h2,x}$ | estimated intercept from univariate LD score regression for the GWAS of the first phenotype |
| $\alpha_{h2,y}$ | estimated intercept for GWAS of the second phenotype |
| $\alpha_{rg}$ | estimated intercept from bivariate LD score regression for the genetic correlation between the two phenotypes |

#### A. Methods

##### *1. Derivation of York regression model with fixed intercept*

We want to compare the trend in genetic effects between the two disorders and identify outliers. To accomplish this we need regression tool allows for (1) sampling errors in GWAS scans for both disease X and Y, and (2) sample overlap, or other sources of correlated sampling errors in the two diseases. York regression is designed for this task, finding a best fit line for two variables that are observed with known measurement error (co)variance. Briefly, for observed variables X and Y, York regression finds the intercept a and slope b such that

$$Y_i = a + bX_i \qquad [1]$$

minimizes the sum of squared weighted residuals

$$S = \sum_{i=1}^{K} \left[\frac{(Y_i - bX_i - a)^2}{\sigma_{y,i}^2 + b^2\sigma_{x,i}^2 - 2b\rho_i\sigma_{x,i}\,\sigma_{y,i}}\right] \qquad [2]$$

where $\sigma_{x,i}^2$, $\sigma_{y,i}^2$, and $\rho_i$ are the squared standard errors and estimated correlation of sampling errors for the two GWAS effect size estimates for each SNP *i*. Compared to standard linear regression, York regression avoids attenuation and inefficient error estimates that would be caused by the ignoring uncertainty in the predictor variable (*X*). This can be thought of as a generalization of Deming regression, which addresses the case where X and Y have error that is independent ($\rho_i = 0$).

For our purposes (see SCOUTJOY below) we want to estimate York regression with the intercept fixed to 0. To our knowledge, no estimator has previously been derived for the fixed intercept case for York regression. However Mahon et al. derived estimators for Deming regression with a fixed intercept, which we closely follow here for generalizing to York regression by allowing for correlated errors[19]. First, let

$$\bar{X} = \frac{\sum_{i=1}^{K} W_i\, X_i}{\sum_{i=1}^{K} W_i} \quad [3]$$

$$\bar{Y} = \frac{\sum_{i=1}^{K} W_i\, Y_i}{\sum_{i=1}^{K} W_i} \quad [4]$$

be the weighted averages across the *K* observations (e.g. loci) normally used in York regression, where *W* is the standard York regression weights for residual variation

$$W_i = \frac{1}{\sigma_{y,i}^2 + b^2\sigma_{x,i}^2 - 2b\rho_i\sigma_{x,i}\sigma_{y,i}} \quad [5]$$

Then as has been shown in Mahon et al., given a fixed intercept $a_0$ the best fit slope can be estimated by:

$$b = \frac{\bar{Y} - a_0}{\bar{X}} \quad [6]$$

As in regular York regression, this estimator for the slope is not a closed form solution since the weights, $W$, used to compute $\bar{Y}$ and $\bar{X}$ depend on the slope, *b*. Instead the slope and weights can be calculated iteratively. The initial slope estimate is made based on ordinary least squares regression with the fixed intercept. $W$, $\bar{X}$, $\bar{Y}$, and *b* are then updated iteratively until *b* converges.

As with previous York papers[17,19], the error variance of the slope can be estimated using the propagation of errors.

$$\sigma_b^2 = \frac{\sum_{i=1}^{k}\left(\frac{\partial\theta}{\partial x_i}\right)^2 \sigma_{x,i}^2 + \left(\frac{\partial\theta}{\partial y_i}\right)^2 \sigma_{y,i}^2 + 2\left(\frac{\partial\theta}{\partial x_i}\right)\left(\frac{\partial\theta}{\partial y_i}\right)\sigma_{xy,i}}{\left(\frac{\partial\theta}{\partial b}\right)^2} \quad [7]$$

Where

$$\theta = a_0 + b\,\bar{X} - \bar{Y} \quad [8]$$

Solving the necessary partial derivatives and substituting $D_i = 2b\sigma_{x,i}^2 - 2\rho_i\sigma_{x,i}\sigma_{y,i}$ yields:

$$\frac{\partial\theta}{\partial x_i} = \frac{bW_i}{\sum_i W_i} \quad [9]$$

$$\frac{\partial\theta}{\partial y_i} = \frac{-W_i}{\sum_i W_i} \quad [10]$$

$$\frac{\partial\theta}{\partial b} = \bar{X} + \frac{\left[(\sum_i W_i)(\sum_i D_i W_i{}^2 Y_i - D_i W_i^2 b X_i)\right] - \left[(\sum_i D_i W_i{}^2)(\sum_i W_i b X_i - W_i Y_i)\right]}{(\sum_i W_i)^2}$$

[ 11 ]

The work above allows us to perform York regression with a fixed intercept. For our application it is also necessary to estimate predicted values for the observed data points based on the fitted regression (sometimes referred to as least squared adjusted points). It can be shown that prior estimators of these predicted values for York regression[17,21] are equivalent[18] and that they still hold with a fixed intercept, which when $a_0 = 0$ simplify to

$$\widehat{X}_i = W_i\left[\left(\sigma_{y,i}^2 - b\rho_i\sigma_{x,i}\sigma_{y,i}\right)X_i + \left(b\sigma_{x,i}^2 - \rho_i\sigma_{x,i}\sigma_{y,i}\right)Y_i\right]$$
$$\widehat{Y}_i = b\widehat{X}_i$$

[ 12 ]

With this we can construct an approach to compare genetic effects in two disorders as per below.

#### 2. Estimation of correlation in sampling error for GWAS effect sizes

York regression assumes that for each observation the sampling errors in $X_i$ and $Y_i$ ($\sigma_{x,i}^2$ and $\sigma_{y,i}^2$, respectively) and their correlation ($\rho_i$) are known. For our application to GWAS loci, standard error estimates for the effect sizes are normally available, but the sampling covariance is not normally estimated. To avoid requiring a joint model to be fit for each locus, we estimate the correlation $\rho_i$ from GWAS summary statistics.

Specifically, it has previously been observed that the covariance of sampling errors in normalized GWAS effect sizes can be estimated, up to scaling factors for sample size, based on the intercept terms of univariate and bivariate LD score regression[22].Thus if $\alpha_{h2,x}$ is the estimated intercept from univariate LD score regression for the GWAS of the first phenotype, $\alpha_{h2,y}$ is the corresponding intercept for GWAS of the second phenotype, and $\alpha_{rg}$ is the estimated intercept from bivariate LD score regression for the genetic correlation between the two phenotypes, then we estimate

$$\rho = \frac{\alpha_{rg}}{\sqrt{\alpha_{h2,x}\alpha_{h2,y}}}$$

[ 13 ]

as the sampling correlation of the GWAS effect sizes for all SNPs in the analysis. We assume this correlation is likely to be stable across SNPs under the assumption that any sampling covariance is likely to be driven by sample overlap and phenotypic correlation and that these values will be constant across SNPs in our current analysis. We therefore use this estimate of $\rho$ for all loci in defining weights for York regression.

#### 3. Significant Cross-trait OUtliers and Trends in JOint York regression (SCOUTJOY)

We are interested in the question of whether there is a consistent relationship in the effect sizes between GDM and T2D or other traits, as might be expected for instance if the loci for GDM reflect exposing the same underlying genetic risk factors as T2D. Genetic correlation estimates can help quantify this relationship on the genome-wide level, but we are interested in also making this comparison among genome-wide significant loci. Our interest is not only in quantifying the average relationship, but in identifying whether the observed locus results are consistent with a uniform relationship between the two phenotypes or if certain loci can be identified as outliers, potentially indicating differential roles or relative importance in each context.

This question is statistically similar to the types of heterogeneity MR-PRESSO[20] was developed to detect in mendelian randomization. Although we explicitly do not intend to draw any causal inferences about the pair of phenotypes, these diagnostic tests match our interest in identifying trends and outliers among the top genome-wide significant loci for a pair of traits.

Therefore we here adapt the tests introduced by MR-PRESSO[20] to our use case, to (a) use York regression to account for measurement error in the effect size estimates from both GWAS, including possible correlated error due to e.g. sample overlap, and (b) extend outlier detect in cases where initial estimates of the trend in correlating effect size estimates may be unstable. We denote this adapted approach as Significant Cross-trait OUtliers and Trends in JOint York regression (SCOUTJOY).

To begin, we identify the lead SNPs for the genome-wide significant loci in the first phenotype (e.g. the 13 top SNPs for GDM) and get their estimated GWAS effect sizes and corresponding standard errors in GWAS of each trait (e.g. GDM and T2D), with effect sizes oriented to the risk increasing allele in the first phenotype. We then use York regression with the intercept fixed to zero (as described above) to estimate the slope representing the average trend in the relationship of the effect sizes for the two phenotypes for the selected SNPs. Fixing the intercept at zero represents a null model that the selected loci based on GWAS of the first phenotype aren't expected to have an average effect on the second phenotype across SNPs that is unrelated to the first phenotype. Adopting this null model facilitates closely following the approach of MR-PRESSO to evaluating excess heterogeneity across SNPs relative to this null trend.

After fitting this initial trend line by York regression, we then evaluate both (1) general overdispersion relative to a single trend line and (2) assess whether individual SNPs can be identified as outliers relative to the effects seen for other SNPs in the group. Outliers are of particular interest, so adaptations were made to extend the current approach of MR-PRESSO.

*i. <u>Evaluation of general overall dispersion using the Global Test</u>*

Our dispersion test is structured analogously to the general approach taken previously for MR-PRESSO[20] with a few changes. This approach evaluates whether the distribution of observed residuals from each point to the fitted regression line are larger than would be expected if the true SNP effect sizes followed a single line. In contrast to MR-PRESSO, we use York regression to fit the regression line which allows for correlated sampling error in both sets of betas in each study under comparison.

More specifically, this is a 4 step process:

(a) For each variant, *i*, we perform a leave-one-out analysis to calculate the regression line using York regression on the remaining variants, $\hat{b}_{-i}$. This is the slope of the trend without point *i*, and allows evaluation of residual variation without overfitting of the slope to the variant.

(b) Based on the estimated trend line, $\hat{b}_{-i}$ we calculate the observed weighted squared residual value (RE) for each of the *K* variants using the York regression weights and sum across SNPs:

$$RE_{OBS} = \sum_{i-1}^{K} RE_{OBS}(i) = \sum_{i-1}^{K} \left[ \frac{\left(Y_i - \hat{b}_{-i} X_i\right)^2}{\sigma_{y,i}^2 + \hat{b}_{-i}^{\ 2} \sigma_{x,i}^2 - 2\hat{b}_{-i}\rho_i \sigma_{x,i}\, \sigma_{y,i}} \right] \quad [14]$$

(c) As in MR-PRESSO, the observed RE is compared against a simulated null distribution for expected REs under the null hypothesis that the true GWAS effect sizes fall on a single trend line. To simulate this null distribution, *M* simulation replicates for each variant is drawn from a bivariate normal distribution:

$$\begin{pmatrix} X_i^{(m)} \\ Y_i^{(m)} \end{pmatrix} \sim N\left( \begin{bmatrix} \widehat{X}_i \\ \widehat{Y}_i \end{bmatrix}, \begin{bmatrix} \sigma_{x,i}^2 & \rho_i \sigma_{x,i}\, \sigma_{y,i} \\ \rho_i \sigma_{x,i}\, \sigma_{y,i} & \sigma_{y,i}^2 \end{bmatrix} \right) \quad [15]$$

where $\widehat{X}_i$ and $\widehat{Y}_i$ are estimated as in Equation 12 using the leave-one-out estimate $\hat{b}_{-i}$. We then compute the weighted squared residuals for each variant in each simulation replicate to obtain a null distribution of *M* expected REs.

$$RE_{EXP}^{m} = \sum_{i} RE_{EXP}^{(m)}(i) = \sum_{i-1}^{K} \left[ \frac{\left(Y_i^{(m)} - \hat{b}_{-i} X_i^{(m)}\right)^2}{\sigma_{y,i}^2 + \hat{b}_{-i}^{\ 2} \sigma_{x,i}^2 - 2\hat{b}_{-i}\rho_i \sigma_{x,i}\,\sigma_{y,i}} \right]$$

*[ 16 ]*

(d) The observed sum of residuals is then compared to the distribution squared residuals from the *M* simulated null replicates to compute an empirical p-value[23] for whether the observed squared residuals are more overdispersed than would be expected:

$$P = \left[ \frac{1 + \sum_{\mathrm{m}} I(RE_{OBS} > RE_{EXP}^{m})}{M+1} \right]$$

*[ 17 ]*

where $I()$ is the indicator function. A significant P value for this test indicates that the observed effect sizes for the two phenotypes are not consistent with a uniform trend in effects across the included SNPs.

*ii. <u>Identification of outliers</u>*

Beyond overall overdispersion, we would like to identify if individual points are outliers relative to the bivariate trend line. Using the observed residuals and simulated null replicates from the Global Test we can use the same strategy as MR-PRESSO to compute the empirical p-value

$$P_i = \left[ \frac{1 + \sum_{\mathrm{m}} I\big(RE_{OBS}(i) > RE_{EXP}^{m}(i)\big)}{M+1} \right]$$

*[ 18 ]*

for each variant to test whether it is an outlier. We evaluate this p-value is evaluated after correction for multiple testing of the *K* variants in the analysis.

This test, however, focuses on whether a given variant is an outlier relative to the leave-one-out estimate of the slope ($\hat{b}_{-i}$). If there are multiple outliers present among the included variants, this means the outlier status of the SNP is compared to a regression that is potentially biased by other outlier points. Given we are specifically interested in identifying outliers that might be scientifically informative we attempt to reduce this potential influence of the presence of multiple outliers by iterating the outlier detection process until the results appear stable.

The first step in outlier identification is directly analogous to what is seen in MR-PRESSO where the observed residual of each point is compared against the residuals of the simulated residual null distribution across all SNPs. However we extend this approach by performing iterative outlier detection against a regression fit that excludes previously identified outliers and repeating this until no outliers remain. We also evaluate whether previously identified outliers are no longer outliers after updates to the regression fit.

Specifically, the iterative outlier detection process works as follows:

1. For each of the K points, test for outlier status by computing $P_i$ from $RE_{OBS}(i)$ and $RE_{EXP}(i)$ based on leave-one-out York regression as described above. Denote the set of variants that are significant after Bonferroni correction for multiple testing as $S_0$. If there are no identified outliers then the procedure stops here.
2. For each of the K points, perform a new leave-*n*-out York regression excluding both the *i*th variant and the set of putative outliers $S_0$. Simulate new null replicates and compute new $P_i$

from observed and simulated residuals for each variant. From these new p-values determine which of the K points are significant outliers after Bonferroni correction for K variants, denoting this new set as $S_1$. Note that this means that an outlier detected in a previous iteration could be returned to "non-outlier" status if it no longer appears to be an outlier after other exclusions.

3. If the putative outlier sets $S_0$ and $S_1$ are identical then the outlier detection has converged. If $S_1$ does not match $S_0$ but is identical to an outlier set identified in some previous iteration then the procedure will become stuck in a loop and so iteration is halted without convergence. Otherwise $S_0$ is updated to match $S_1$ and step 2 is repeated.

If the process converges then the variants in the final stable set $S_1$ are concluded to be significant outliers relative to the primary trend of relationship in GWAS effect sizes between the two phenotypes. While convergence is not guaranteed, in practice we do not encounter issues with infinite loops in the current analysis.

**B. Results**

For the 13 GDM-associated loci, we compared the genetic effects in GDM with 5 other disorders including Type 2 diabetes, fasting glucose, HbA1C, 2h Glucose on Oral Glucose Tolerance Testing (OGTT) and fasting insulin. The comparison to T2D showed a group of outliers with strong T2D effect indicating a stratification that was not seen in any of the other glycemic traits, (Supplementary Figure 22). Falling into more than one group suggests some variants might have a different effect in pregnancy than the majority of variants that have an effect in both GDM and T2D. This would be further explored in the shared variants analysis. After removal of the outliers, there was a significant positive relationship between genetic effects in GDM and in T2D for the 13 loci (Supplementary Table 21).

While the other glycemic traits did not have a stratified result seen in T2D, the relationship between genetic effects in GDM and in other traits (the trend line slope) was still informative (Supplementary Figure 21, Supplementary Table 20). A significant positive relationship was seen in fasting glucose (slope 0.19, p-value =5.2e-12) , HbA1C (slope 0.07, p-value =2.35e-10), and 2h glucose on OGTT(slope 0.15, p-value =4.0e-6) which suggests a uniform expected positive effect in SNPS in both disorders. In fasting insulin, no significant relationship was seen (slope= -1.24E-03, p-value=0.79). Complete list of slopes in Supplementary Table 20.

**C. Discussion**

We were able to compare genetic effects across related diseases using our new method, SCOUTJOY, that leverages a newly derived regression form. This detected a unique relationship between GDM and T2D that was not present with other traits and could detail the relationship or lack thereof with glycemic traits.

### VI. Genetic effect differences by sex or pregnancy history

GDM is only experiences by pregnant females, so any observed differences between genetic effects for GDM and T2D could be confounded with differences in T2D genetic effects by sex or pregnancy history. We therefore evaluated whether the observed relationship of genetic effects between GDM and T2D was consistent across T2D in different groups by sex or pregnancy history.

We first performed a genome wide association for T2D in (a) T2D in males, (b) T2D in females, (c) T2D in parous females who had a history of pregnancy and (d) T2D in nulliparous females with no history of pregnancy. Within each group, cases were those with T2D and controls were those without T2D.

The stratified T2D GWAS allow comparison of sex or pregnancy effects in several ways. First, we test each of the 13 top hits from GDM for differences in T2D effect sizes between males and females or between parous and nulliparous females (method below). Second, we assess whether the overall trend in effect sizes for GDM and T2D across the 13 GDM hits differs when comparing to T2D in males vs. females or parous vs. nulliparous females using SCOUTJOY. Third, we consider the genome-wide genetic correlation of GDM with T2D in each sex and pregnancy history groups, as well as genetic correlation of T2D GWAS between the different groups.

#### A. Method for comparing genetic effects of a SNP in two traits

To determine if any of the GDM hits showed differences in T2D effects by sex or pregnancy history we compared the effect of each locus between groups. Following the same method previously used for testing sex differences in GWAS effect sizes[24], for each comparison (i.e. male vs. female, parous vs. nonparous) we computed the Z-score for the difference in observed genetic effects in each of two nonoverlapping groups for each top GDM SNP *i*:

$$Z_i = \frac{\beta_{1,i} - \beta_{2,i}}{\sqrt{se_{1,i}^2 + se_{2,i}^2}}$$

P-values for the comparison for each SNP are obtained from a two-sided test of this Z-score, and evaluated after Bonferroni correction for multiple testing of the 13 GDM loci.

#### B. Results

For the 13 GDM-associated loci, we compared the genetic effect on T2D in men versus women. Direct comparison of genetic effects at these loci revealed a significant difference at the CKDN2B locus (Supplementary Table 24). We then evaluated the relationship of genetic effects in T2D in men versus women using SCOUTJOY, (Supplementary Figure 23, Supplementary Table 21). The magnitude of genetic effect were very similar (slope=1.00, se=0.06) between sexes after identification of the one outlier at the CDKN2B locus. Effect plots of GDM versus T2D in each sex generally concordant, (Supplementary Figure 23) with the broadly similar slopes for the relationship of GDM with T2D in males (slope=0.44, se=0.05) and with T2D in females (slope=0.31, se=0.03).

Comparison of the genetic effects in T2D was then performed for females with a history of pregnancy (parous females) and those without (nulliparous females). Direct comparison revealed that none of the 13 GDM-associated loci were significantly different in these groups (Supplementary Table 24). Evaluation of the relationship of genetic effects using SCOUTJOY (Supplementary Figure 24, Supplementary Table 21) similarly revealed no outliers or heterogeneity (global test p=0.41) and generally concordant effect sizes (slope=0.77, se=0.11), though with higher uncertainty in nulliparous females due to lower sample size.

We further assessed genome-wide genetic correlation of GDM with T2D in men and women with different pregnancy histories, respectively (Supplementary Table 23). Highly similar genetic correlations were observed for GDM with males ($r_g$ =0.76, se = 0.062), females ($r_g$ =0.78, se =0.067), and parous females ($r_g$ =0.77, se =0.074). While the correlation of GDM vs nulliparous females ($r_g$=0.94, se=0.16) appears different from the GDM vs the other subgroups, there's no sign that the genetics of T2D in parous females and nulliparous females actually differs ($r_g$=1.18, se=0.16). The initial observation could be attributable to sampling variation given the wider standard error and smaller sample size of the GWAS in nulliparous females. We do however find evidence that T2D in males is not perfectly genetically correlated with T2D in parous females (Z-score 1.73, p = 0.01) despite the high genetic correlation ($r_g$ 0.92), potentially consistent with the observation of a single outlier with sex differences at the locus level for CDKN2B.

#### VII. Selection of variants for shared variants analysis

As stated in the Online Methods, we used the linemodels package (https://github.com/mjpirinen/linemodels) to compare summary statistics for 28 lead variant from both the T2D and GDM GWAS (13 from GDM and 15 from T2D).

Our goal was a fair comparison of effect sizes between our significant GDM loci and previously known T2D loci we might expect to have similar power to detect under a null hypothesis that the effect size in GDM is the same as in T2D. T2D has a higher effective sample size in Finngen, because it is both more common and is not restricted to parous females. Thus to balance the comparison, we took only the T2D loci that would have been identified in a smaller study equivalent to the power of the GDM GWAS.

To do this, we first identified the minimum standardized effect size $\beta^*_{min}$ among the genome-wide significant GDM loci (i.e. standardized for allele frequency).

$$\beta_j^* = \beta_j \sqrt{(2p_j(1 - p_j)}$$

$$\beta^*_{min} = \min_j \left|\beta_j^*\right|$$

where $\beta_j$ is the estimated log odds ratio for variant *j* in GDM GWAS, $p_j$ is the minor allele frequency for variant *j* and $\beta_j^*$ is the standardized effect size for variant *j*. It can be shown that the expected GWAS $\chi^2$ statistic for a variant with observed standardized effect size $\beta^*$ can then be approximated by

$$\chi_j^2 \approx NK(1 - K){\beta_j^*}^2$$

where *N* is the GWAS sample size and *K* is the proportion of cases in the GWAS sample. It may be noted that $NK(1 - K)$ is proportional to conventional estimates of effective sample size. We therefore estimate that a variant with the minimum detected standardized beta from the GDM GWAS $\beta^*_{min}$ would correspond to an expected GWAS $\chi^2$ of

$$\chi^2_{j,T2D} = N_{T2D}K_{T2D}(1 - K_{T2D}){\beta^*_{min}}^2$$

in the Finngen GWAS of T2D with sample size $N_{T2D}$ and in-sample prevalence $K_{T2D}$.

Based on the sample size for T2D in Finngen release 6 (37031 cases, 214308 controls) we find the resulting $\chi^2_{j,T2D}$ corresponds to a p-value of 1.27e-22. Using this threshold we identified 15 independent loci from the T2D GWAS passing this p-value threshold that we would therefore expect to have power to detect in the GDM GWAS if the GDM and T2D effect sizes were equal. We therefore use the lead variants for this set of 15 T2D loci along with the 13 significant GDM loci for our comparison of top hit effect sizes.

### VIII. Comparison of relationship of genetic effects in BMI and GDM for Class T and Class G loci

After identifying Class T and Class G SNPs, we were interested in their potential relationship with BMI. We focus here on BMI because we are not well powered to evaluate apparent patterns in other glycemic traits due to the smaller sample size of their published GWAS. For each class we compared the effects of member loci in gestational diabetes vs BMI using SCOUTJOY (Supplementary Figure 26, Supplementary Table 24). For each of the classes of SNPs we calculate the relationship between SNP effect in GDM and SNP effect in BMI IRN. We then compare the relationships observed across classes.

#### A. Method to generate and compare genetic effect relationships

Summary statistics from the GWAS for GDM, T2D, and inverse rank normalized BMI (BMI IRN) were used from Finngen release 8. We then compared the effects using SCOUTJOY algorithm as detailed above. Comparison of slopes from these independent sets of variants was performed using Z-score.

$$Z = \frac{slope_{Class_T} - slope_{Class_G}}{\sqrt[2]{se(slope)^2_{Class_T} + se(slope)^2_{Class_G}}}$$

#### B. Results

The Class G loci, which have a GDM-predominant effect, had no significant relationship between the genetic effect in GDM and in BMI IRN across all variants (slope=0.009, p=0.22; Supplementary Figure 26a, Supplementary Table 24), and only a weak relationship after exclusion of 1 outlier (slope=0.026, p=0.010). In contrast, the Class T loci, which have predominant effect in T2D, had a significant relationship between the genetic effect in GDM and BMI after outlier removal (beta -0.12, p 6.8e-07). The relationship observed between GDM and BMI IRN was significantly different between Class G and Class T loci when the slopes were compared after excluding outliers (Z-score 9.6, p=6.0e-22). The negative relationship seen between top hits in Class T and BMI IRN echoes observations made previously about a negative correlation between genetic effects of top T2D hits and BMI[25]. We observe the same pattern in SCOUTJOY comparison of genetic effect in BMI IRN with T2D for class T loci (Supplementary Figure 26c; slope -0.9, p-value =1.55e-07).

### IX. Cell type specificity analyses

We were interested assessing the relationship between the GDM summary statistics and single cell expression across the body – particularly in comparison to the T2D summary statistics. To that end, we assessed the relationship between genetic summary statistics and single cell RNA (scRNA) datasets to detect relationships with specific cells and tissues. We focused on independent signals from each tissue and then evaluated cross-tissue signal dependence related to each disease. We performed this analysis on cell-specific tissue expression despite the lack of significant findings on bulk level tissue expression (Supplementary Figure 16 and 17) because we know that adaptive physiologic changes in pregnancy induces major adaptive changes to specific cell populations within maternal tissues and effects on expression specific to these cell populations might be obscured in bulk tissue expression.

#### A. Datasets for analysis

Genetic datasets tested include (1) our GDM summary statistics and, for reference, for (2) summary statistics from a recent Type 2 Diabetes European meta analysis dataset[26].

The scRNA datasets were tested in a staged manner, summarized below. We first performed this analysis on a large, well curated murine scRNA dataset that provides survey level data across 22 different organ/tissue types. Based on the results of this analysis, significant tissues were then more carefully evaluated with tissue specific murine datasets from hypothesized brain regions of interest. In the tissues/organs indicated, we also ran analyses on high quality human scRNA datasets where possible and provide comparisons.

| Name | Description | PMID column |
|---|---|---|
| Tabula Muris FACS | Mouse samples were tested resulting in 119 cell-tissue pairs from 22 tissues/organs. scRNA expression was measured in 53,760 cells in the raw read count matrix, after QC 44,949 cells exist in the annotation file. Cells with label "unknown" were included as it was stated in the original study that they are potential novel cell types. Number of genes tested was 23,433 genes with 15,131 genes were mapped to human ENSG ID. | 30283141 (ref.[27]) |
| Mouse Hypothalamus (GSE87544) | Mouse hypothalamus samples were obtained from 8-10 week old mice and tested resulting in 46 cell types from hypothalamic tissue. scRNA expression was measured in 14,437 cells, after QC the total number of cells used was 1,039. Number of genes tested was 23,284 genes with 15,116 genes were mapped to human ENSG ID. | 28355573 (ref.[28]) |
| Mouse arcuate ME neurons (GSE93374) | Mouse brain samples were obtained from the hypothalamic arcuate-median eminence complex in 4-12 week old mice. Testing resulted in 28 cell types from the arcuate nucleus tissue. scRNA expression was measured in 21,086 cells resulting in 13,079 cells after QC and clustering. Number of genes tested was 19,743 genes of which 14,366 genes were mapped to human ENSG ID. | 28166221 (ref.[29] ) |
| GSE84133 | Mouse: All 1,886 cells were and from 14,878 genes, 12,741 genes were mapped to unique human ENSG ID. | 27667365 (ref[30]) |
| GSE84133 | Human pancreas samples from individuals without diabetes. 7,266 cell were sampled. Number of genes tested was 20,125 genes and 19,546 genes were mapped to unique ENSG ID. | 27667365 (ref[30]) |

For complete dataset details see summarized descriptions (https://fuma.ctglab.nl/tutorial#celltype). Multiple testing correction was performed using the Benjamini-Hochberg multiple test correction (FDR).

#### B. Method for cell specificity analysis

Assessing relationships across multiple scRNA-seq datasets is made particularly challenging due to complex batch and sampling differences between studies. A method addressing these concerns has been described by Wantanabe et al[31]. This method for a cell specificity analysis has been made available as part for the FUMA package. We briefly review the process below but for details please refer to the paper and online tutorials (https://fuma.ctglab.nl/tutorial#celltype). The analysis is a three-step process.

##### *1. Identify cells with scRNA expression significantly associated with genetic summary statistics*

For each cell/tissue type in each dataset, gene-level GWAS summary statistics from MAGMA are regressed on gene expression levels for the cell/tissue type and technical covariates.

##### *2. Identify independently associated cell types within each dataset*

For each dataset, all pairs of cell/tissue types that were significantly associated with the GWAS MAGMA results in Step 1 are systematically tested for conditional association. For each cell type pair, a proportional significance (PS) is then calculated:

$$PS_{\tau,\varphi} = \frac{-\log_{10}(\mathrm{p}_{\tau,\varphi})}{-\log_{10}(\mathrm{p}_{\tau})}$$

Where $\mathrm{p}_{\tau}$ is the marginal p-value for a cell type and $\mathrm{p}_{\tau,\varphi}$ is the conditional p-value for cell type $\boldsymbol{\tau}$ conditioning on cell type $\boldsymbol{\varphi}$. Using the PS score for each cell type, the signal between cell type is then determined to be dependent or independent.

##### *3. Identify independent signals across cells in different datasets/tissues*

All cell/tissue types showing within-dataset conditional association in Step 2 are compared across datasets to identify cell/tissue types that have substantial independent association (PS > 0.5) with the GWAS MAGMA results.

#### C. Results

First we performed the initial step of cell specificity analysis on the summary statistics for GDM on single cell RNA expression data from the Tabula Muris dataset which gives a high-level survey of expression data across 22 tissues in a well-characterized murine sample (Supplementary Figure 26, Supplementary Table 26). Differences were seen between GDM and T2D in this initial screen with GDM having (1) significant associations with cells in the brain not seen in T2D, and (2) different pancreatic cell types being associated with GDM than T2D. Because the cell-level brain data in Tabula Muris was limited, we then repeated the step 1 analysis augmenting the Tabula Muris dataset with data from several high quality scRNA studies of the murine brain. From this analysis, differences in independent signals in cell-tissue data between GDM and T2D were seen (Figure 3, Supplementary Table 27) that reveal GDM was associated with two cell populations in the hypothalamus and one in the arcuate nucleus that were distinct from the one cell-type association in the arcuate nucleus seen in T2D. This is of particular interest as it is well established that adaptive changes in pregnancy occur in the ventromedial hypothalamus (VMH) and arcuate nucleus to regulate blood sugar and blood pressure during pregnancy[32].

To consider the relationship signal in the cells of the brain had with cells in the pancreas, we performed cross dataset conditional analyses on these data (Supplementary Figure 27, Supplementary Table 28) which revealed that one of the GDM hypothalamic cell associations in Glu7 appeared related to the arcuate nucleus cell association Nr5a1-Adcyap1. In the initial paper describing the arcuate nucleus dataset[29], the Nr5a1-Adcyap1 cell population was described as arising from the VMH and projecting across the arcuate nucleus, raising the hypothesis that this could be a direct shared signal. Cells arising from the VMH and projecting across the arcuate nucleus have been shown to express estrogen receptor alpha (ERα) and control glucose balance[33] and potentially be regulated by estrogen levels.[34] The gene encoding ERα, ESR1, is one of our GWAS hits for GDM, and estrogen is known to be elevated significantly in pregnancy.

The conditional analysis of cell specificity also finds that the significantly associated pancreatic cells represent two signals but with some signal overlap (Supplementary Figure 28, Supplementary Tables 28 & 29). The appearance of both murine beta cells and ductal cells as independent but related signals is interesting given ongoing recent work elucidating the role of ductal cells in murine beta cell compensation or expansion during pregnancy[35,36]. Signal in the pancreatic cell types appear distinct from that in the cells of the hypothalamus or arcuate nucelii and in fact that the PS > 1 implies that these signals appear stronger after accounting for signals in the brain (and vice versa).

Finally, the structure of the mouse pancreas has known anatomic, physiologic, and molecular differences from the human pancreas. To compare the relationship of GDM summary statistics with human vs murine pancreatic expression, we performed the initial cell-specificity analysis for both human and mouse pancreatic data (Supplementary Figure 29, Supplementary Table 29). Significant differences in the cell-tissue relationships were seen in human vs murine scRNA expression. GDM was primarily associated with beta cells in the mouse but delta cells in human studies. This analysis of pancreatic cells alone does not show the association with ductal cells in mouse observed in analysis with the full 22 tissue Tabula Muris data, possibly indicating that some of that signal is associated with average expression across pancreatic cell types. In either case, larger human scRNA studies and additional characterization of the signal in mouse cells will be required to elucidate how the relevant aspects of GDM genetics manifest at the cellular level in the pancreas in mice and humans.

### Supplementary Figures

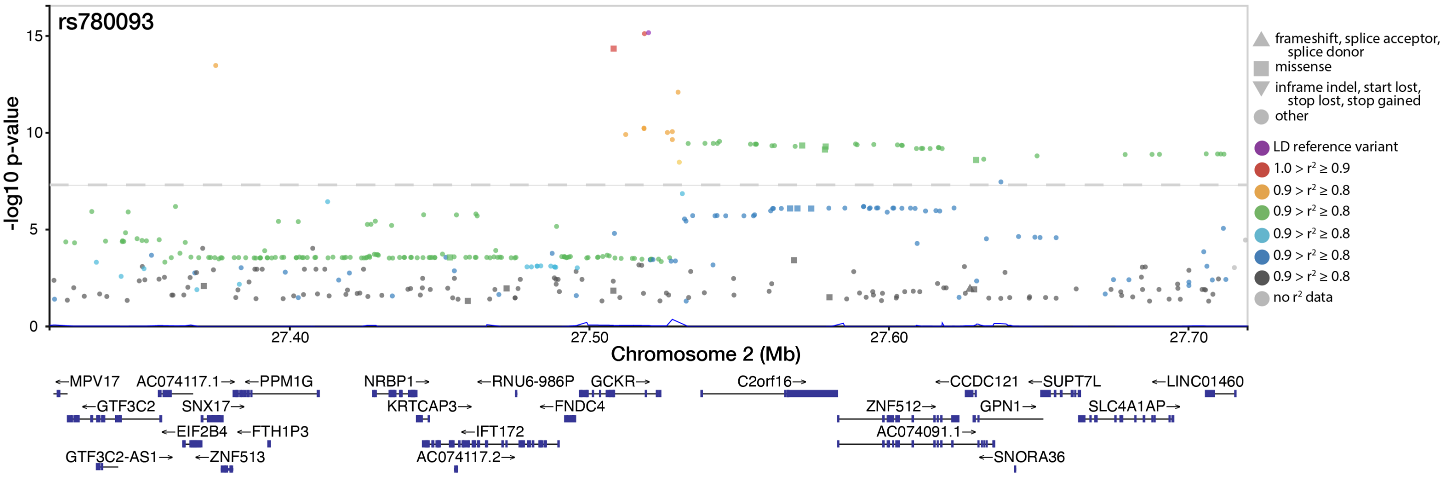

**Supplementary Figure 1: Regional GWAS results for rs780093 locus on chromosome 2 at 27519736.**
SNP label is the lead posterior inclusion probability (PIP) SNP from fine mapping. Dashed reference line is Bonferroni significance threshold (p = 5E-08). Color indicates LD with the lowest p-value SNP in the region. LD is computed from the Finnish SISu v3 reference panel. Shape indicates annotated functional consequence. Genes in the region from GENCODE are annotated below the X-axis.

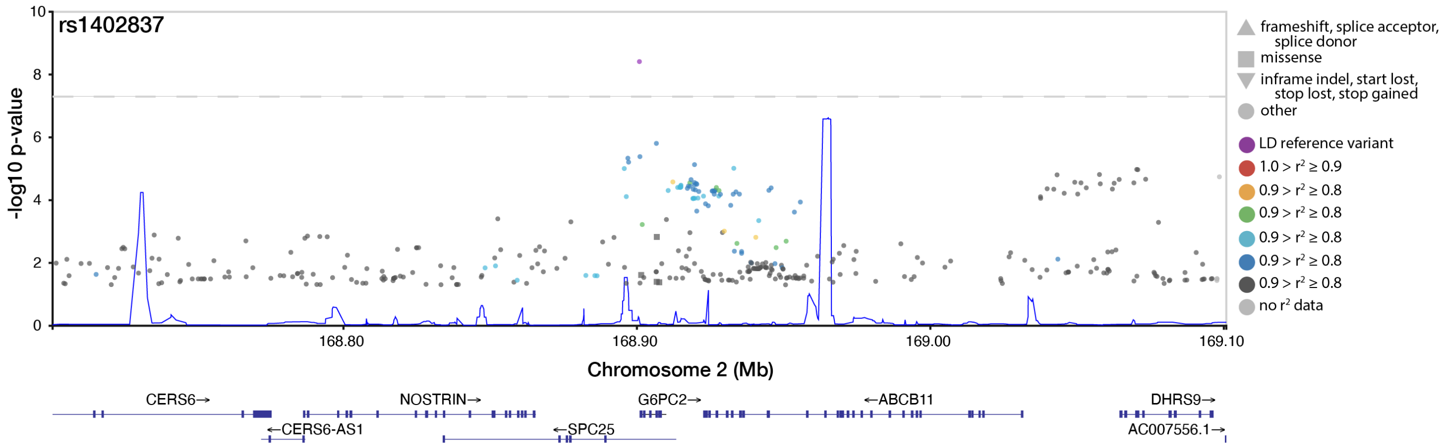

**Supplementary Figure 2: Regional GWAS results for rs1402837 locus on chromosome 2 at 168900844.**
SNP label is the lead posterior inclusion probability (PIP) SNP from fine mapping. Dashed reference line is Bonferroni significance threshold (p = 5E-08). Color indicates LD with the lowest p-value SNP in the region. LD is computed from the Finnish SISu v3 reference panel. Shape indicates annotated functional consequence. Genes in the region from GENCODE are annotated below the X-axis.

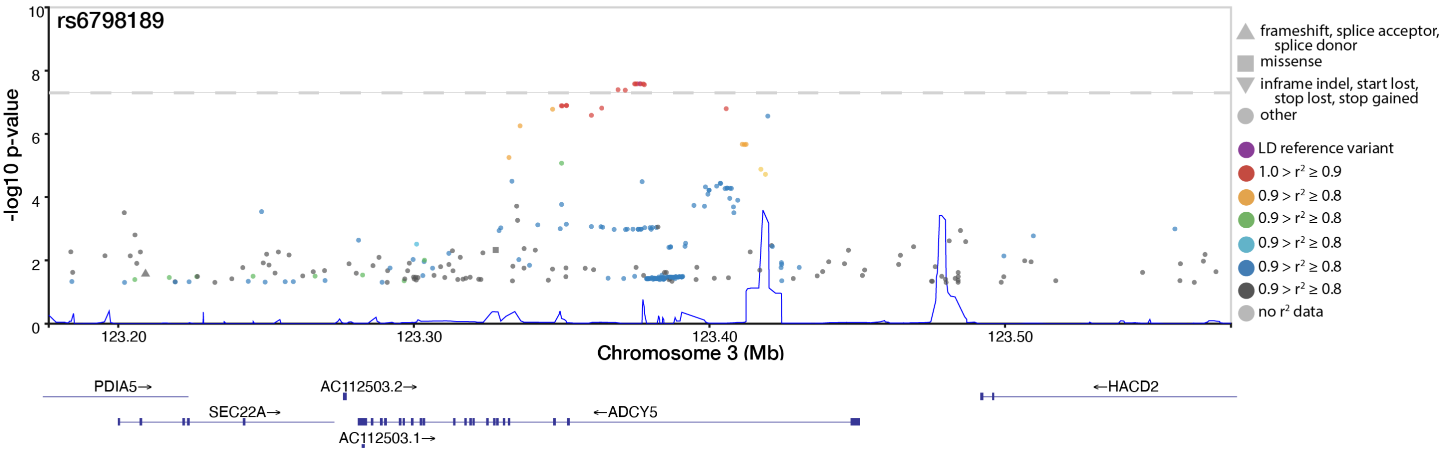

**Supplementary Figure 3: Regional GWAS results for rs6798189 locus on chromosome 3 at 123376465.**
SNP label is the lead posterior inclusion probability (PIP) SNP from fine mapping. Dashed reference line is Bonferroni significance threshold (p = 5E-08). Color indicates LD with the lowest p-value SNP in the region. LD is computed from the Finnish SISu v3 reference panel. Shape indicates annotated functional consequence. Genes in the region from GENCODE are annotated below the X-axis.

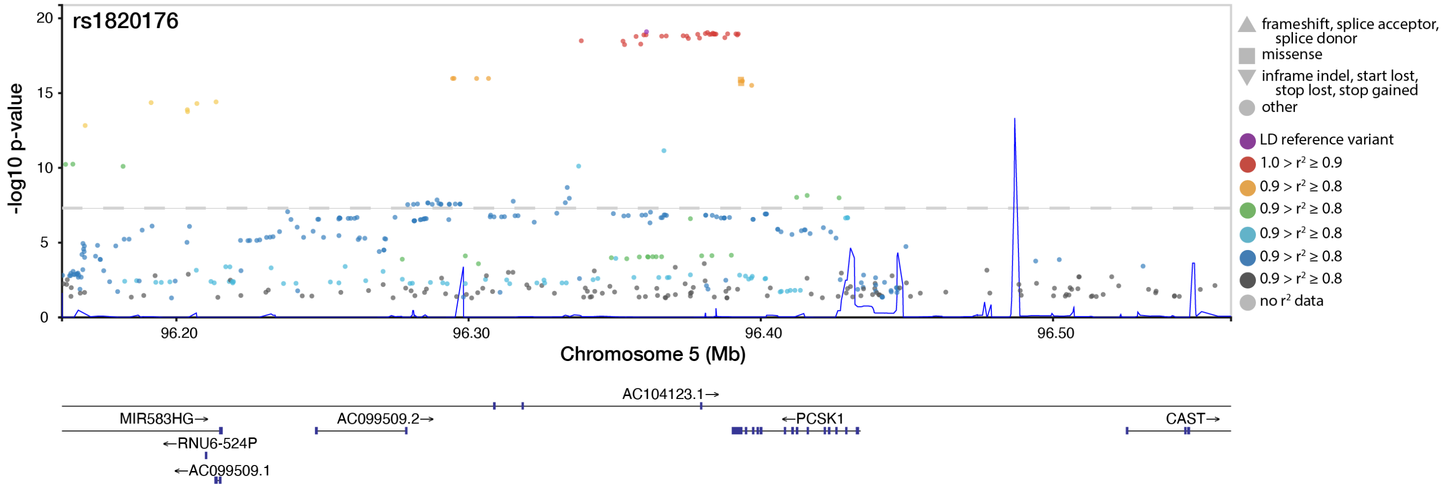

**Supplementary Figure 4: Regional GWAS results for rs1820176 locus on chromosome 5 at 96360881.**
SNP label is the lead posterior inclusion probability (PIP) SNP from fine mapping. Dashed reference line is Bonferroni significance threshold (p = 5E-08). Color indicates LD with the lowest p-value SNP in the region. LD is computed from the Finnish SISu v3 reference panel. Shape indicates annotated functional consequence. Genes in the region from GENCODE are annotated below the X-axis.

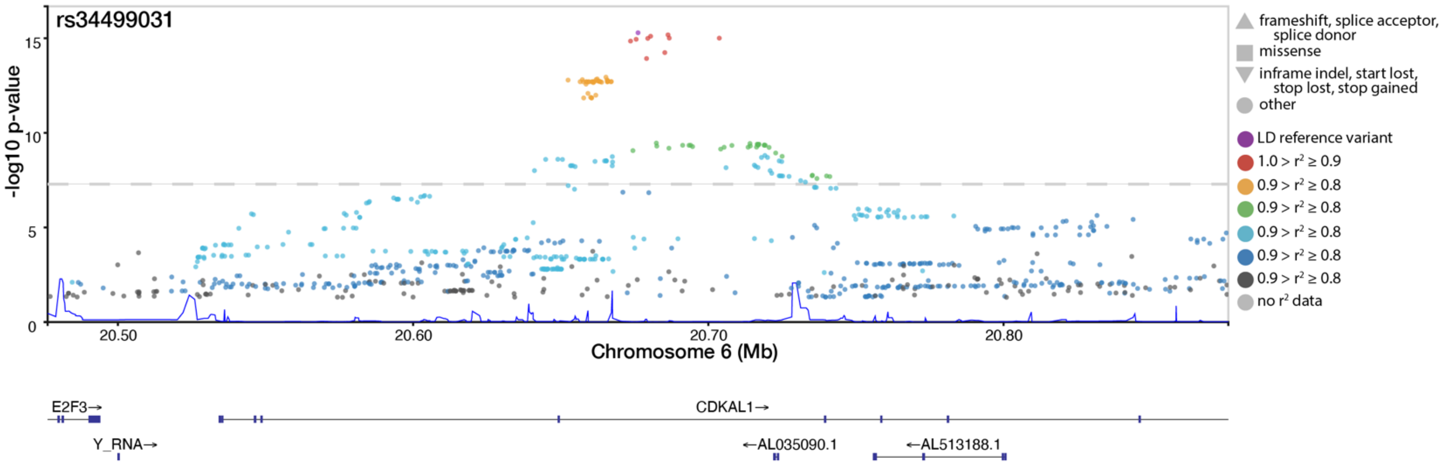

**Supplementary Figure 5: Regional GWAS results for rs34499031 locus on chromosome 6 at 20676183.**
SNP label is the lead posterior inclusion probability (PIP) SNP from fine mapping. Dashed reference line is Bonferroni significance threshold (p = 5E-08). Color indicates LD with the lowest p-value SNP in the region. LD is computed from the Finnish SISu v3 reference panel. Shape indicates annotated functional consequence. Genes in the region from GENCODE are annotated below the X-axis.

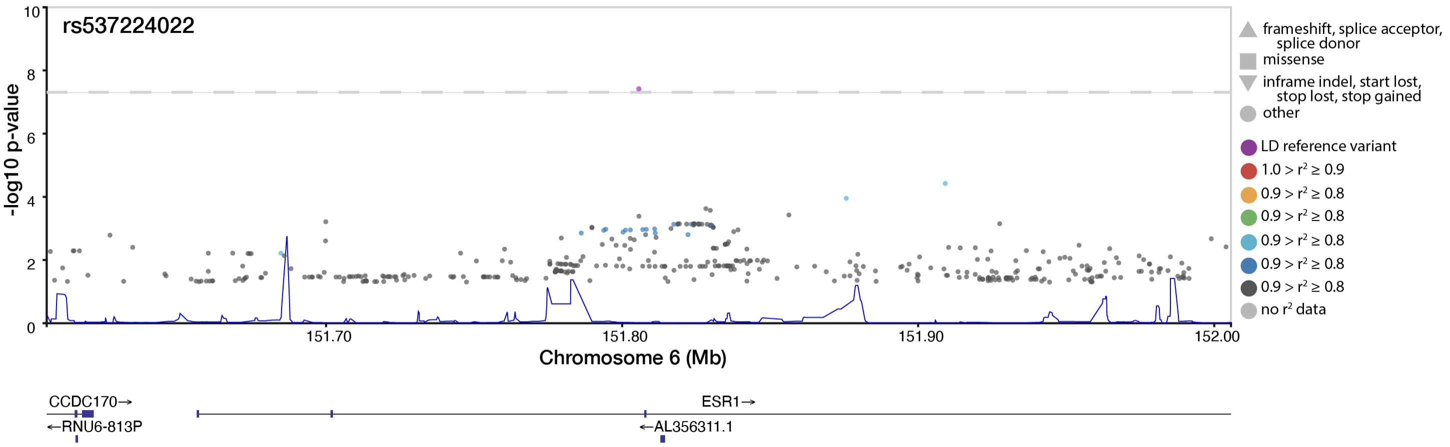

**Supplementary Figure 6: Regional GWAS results for rs537224022 locus on chromosome 6 at 151805650.**
SNP label is the lead posterior inclusion probability (PIP) SNP from fine mapping. Dashed reference line is Bonferroni significance threshold (p = 5E-08). Color indicates LD with the lowest p-value SNP in the region. LD is computed from the Finnish SISu v3 reference panel. Shape indicates annotated functional consequence. Genes in the region from GENCODE are annotated below the X-axis.

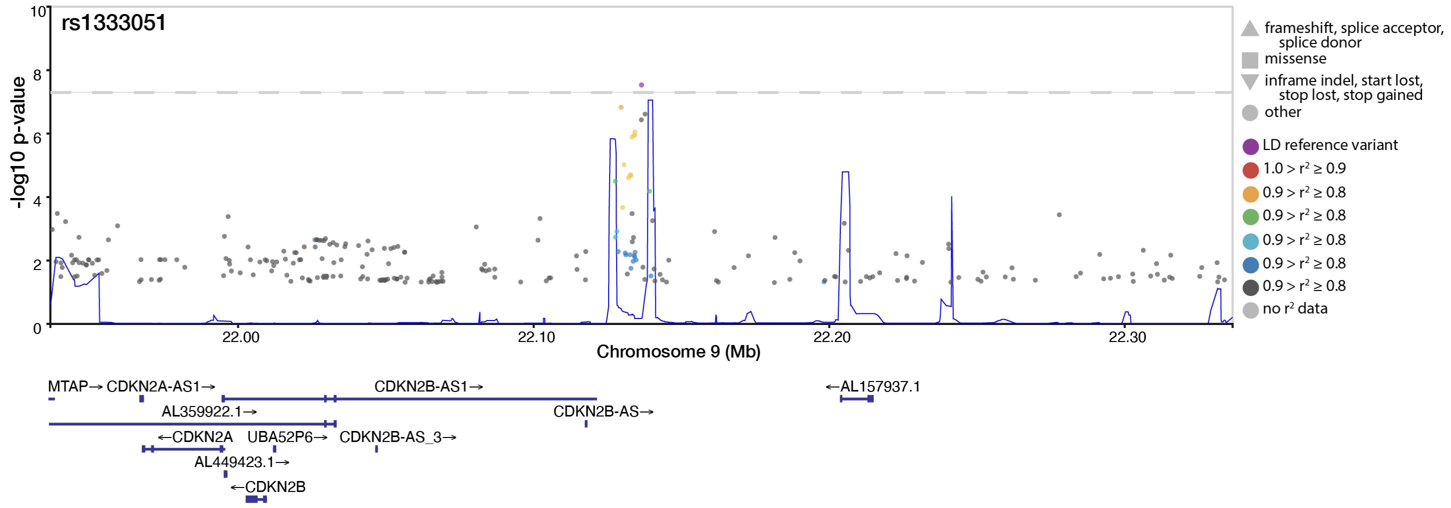

**Supplementary Figure 7: Regional GWAS results for rs1333051 locus on chromosome 9 at 22136490.**
SNP label is the lead posterior inclusion probability (PIP) SNP from fine mapping. Dashed reference line is Bonferroni significance threshold (p = 5E-08). Color indicates LD with the lowest p-value SNP in the region. LD is computed from the Finnish SISu v3 reference panel. Shape indicates annotated functional consequence. Genes in the region from GENCODE are annotated below the X-axis.

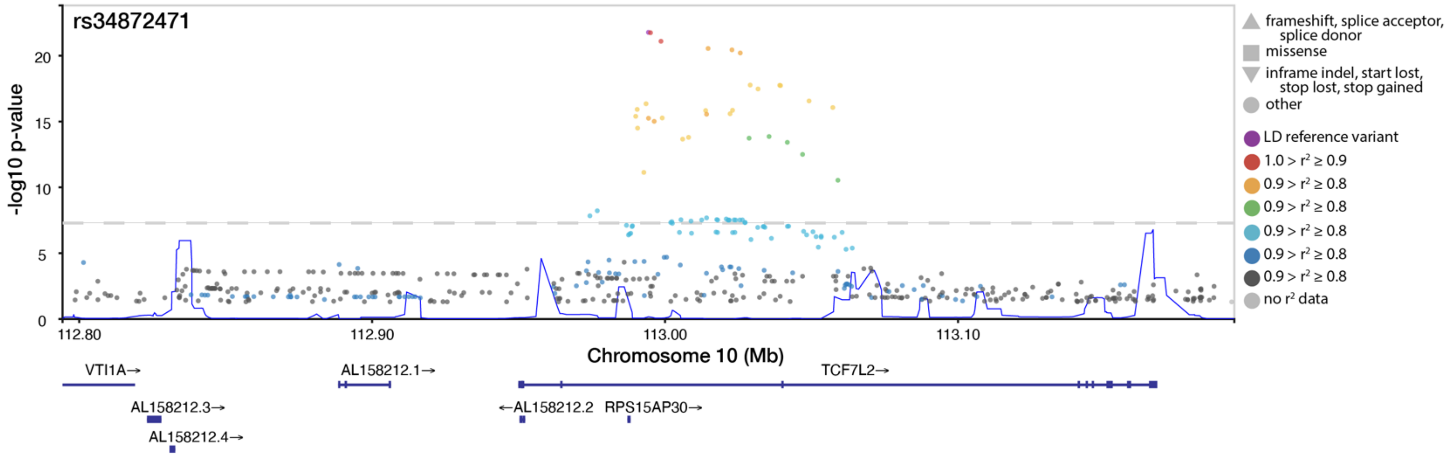

**Supplementary Figure 8: Regional GWAS results for rs34872471 locus on chromosome 10 at 112994312.**
SNP label is the lead posterior inclusion probability (PIP) SNP from fine mapping. Dashed reference line is Bonferroni significance threshold (p = 5E-08). Color indicates LD with the lowest p-value SNP in the region. LD is computed from the Finnish SISu v3 reference panel. Shape indicates annotated functional consequence. Genes in the region from GENCODE are annotated below the X-axis.

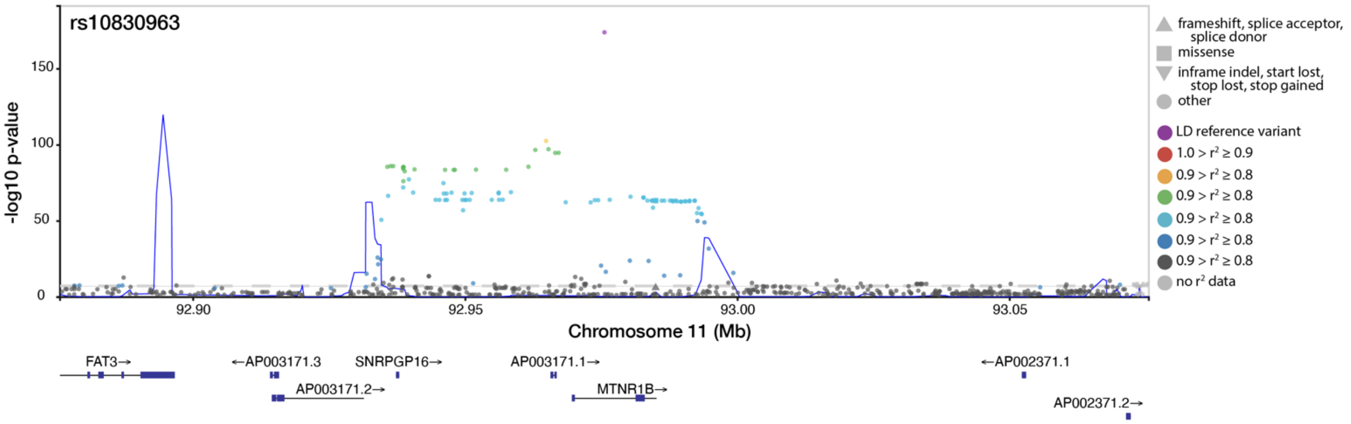

**Supplementary Figure 9: Regional GWAS results for rs10830963 locus on chromosome 11 at 92975544.**
SNP label is the lead posterior inclusion probability (PIP) SNP from fine mapping. Dashed reference line is Bonferroni significance threshold (p = 5E-08). Color indicates LD with the lowest p-value SNP in the region. LD is computed from the Finnish SISu v3 reference panel. Shape indicates annotated functional consequence. Genes in the region from GENCODE are annotated below the X-axis.

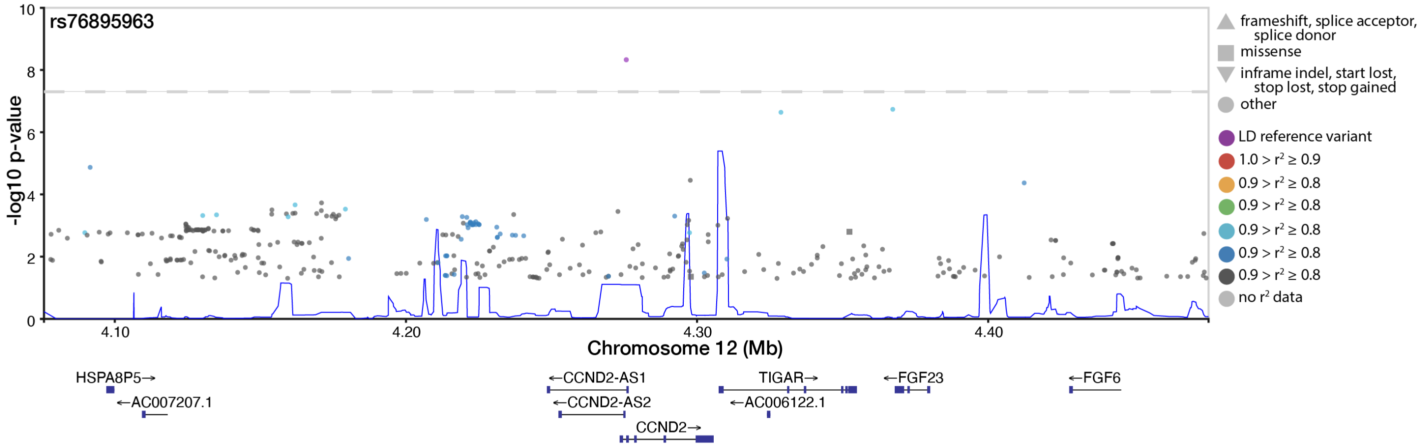

**Supplementary Figure 10: Regional GWAS results for rs76895963 locus on chromosome 12 at 4275678.**
SNP label is the lead posterior inclusion probability (PIP) SNP from fine mapping. Dashed reference line is Bonferroni significance threshold (p = 5E-08). Color indicates LD with the lowest p-value SNP in the region. LD is computed from the Finnish SISu v3 reference panel. Shape indicates annotated functional consequence. Genes in the region from GENCODE are annotated below the X-axis.

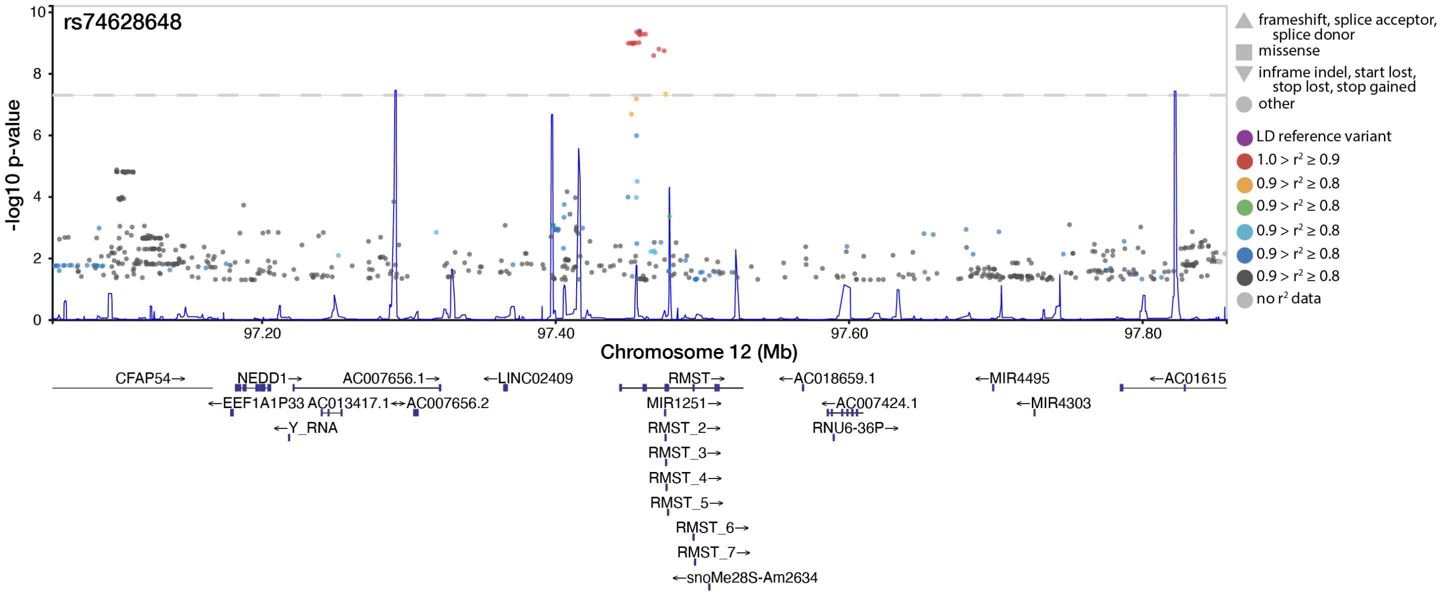

**Supplementary Figure 11: Regional GWAS results for rs74628648 locus on chromosome 12 at 97457224.**

SNP label is the lead posterior inclusion probability (PIP) SNP from fine mapping. Dashed reference line is Bonferroni significance threshold (p = 5E-08). Color indicates LD with the lowest p-value SNP in the region. LD is computed from the Finnish SISu v3 reference panel. Shape indicates annotated functional consequence. Genes in the region from GENCODE are annotated below the X-axis.

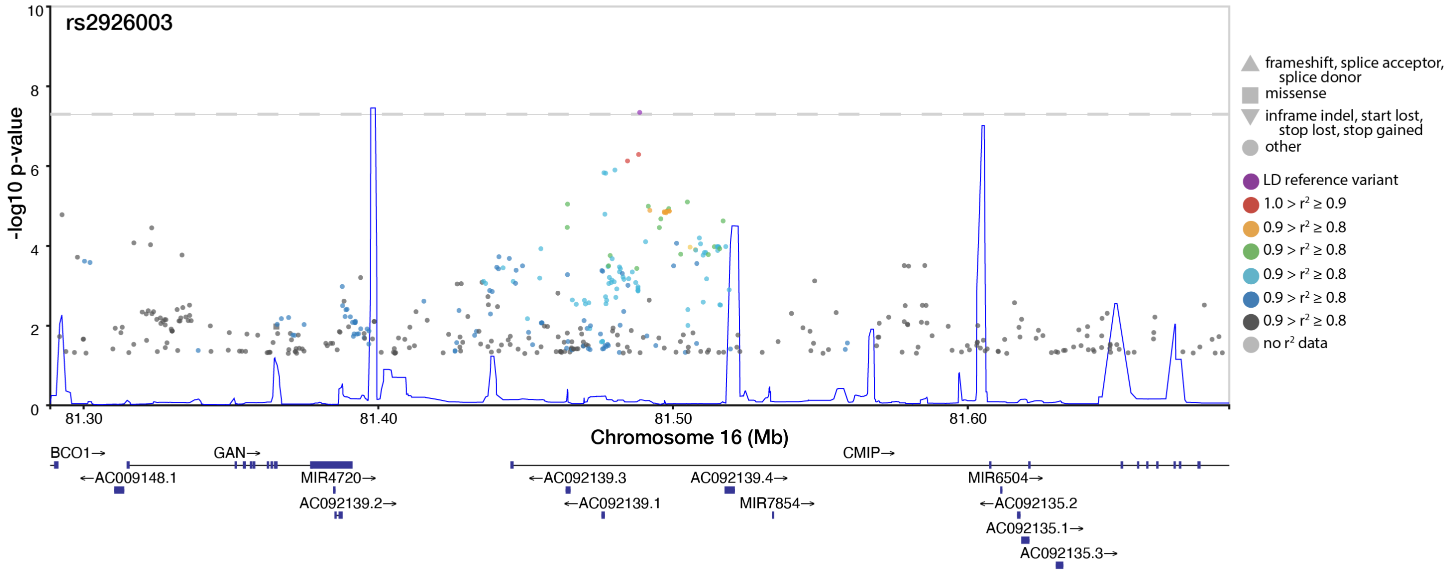

**Supplementary Figure 12: Regional GWAS results for rs2926003 locus on chromosome 16 at 81488676.**

SNP label is the lead posterior inclusion probability (PIP) SNP from fine mapping. Dashed reference line is Bonferroni significance threshold (p = 5E-08). Color indicates LD with the lowest p-value SNP in the region. LD is computed from the Finnish SISu v3 reference panel. Shape indicates annotated functional consequence. Genes in the region from GENCODE are annotated below the X-axis.

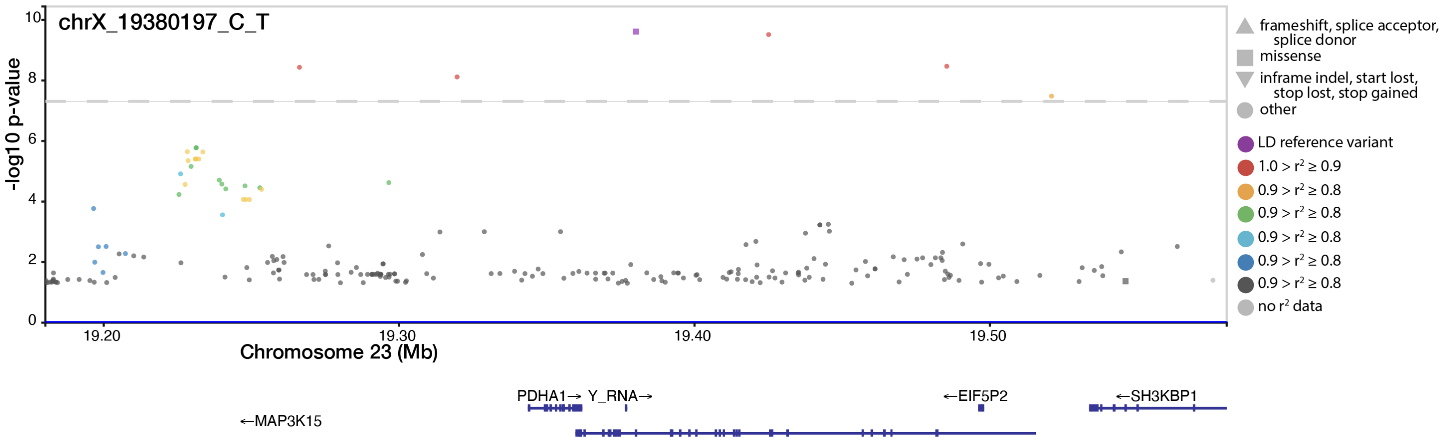

**Supplementary Figure 13: Regional GWAS results for rs56381411 locus on chromosome 23 at 19380197.**

SNP label is the lead posterior inclusion probability (PIP) SNP from fine mapping. Dashed reference line is Bonferroni significance threshold (p = 5E-08). Color indicates LD with the lowest p-value SNP in the region. LD is computed from the Finnish SISu v3 reference panel. Shape indicates annotated functional consequence. Genes in the region from GENCODE are annotated below the X-axis.

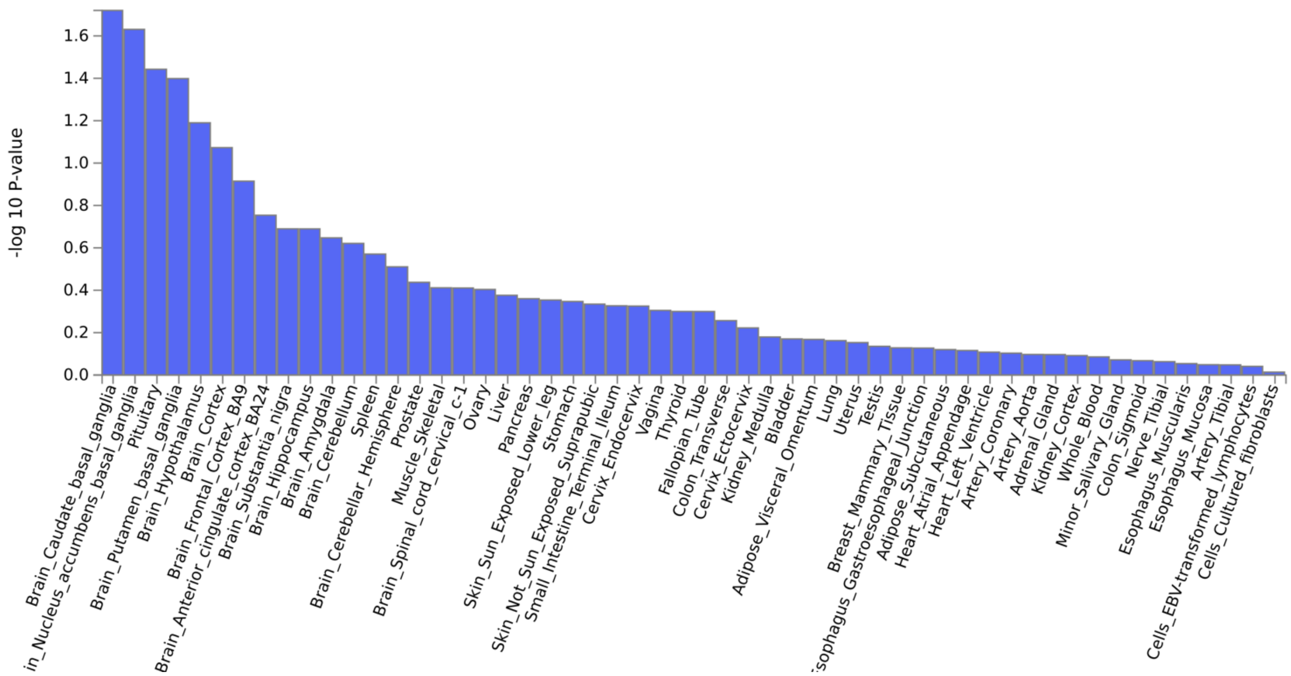

**Supplementary Figure 14: Gene property analysis of tissue specific expression.**
Unadjusted p-values for association of MAGMA gene-level results for GDM with tissue specific gene expression across 17310 genes. Performed using FUMA SNP2GENE based on tissue specificity analysis of expression in GTEx. Data available in Supplementary Table 12.

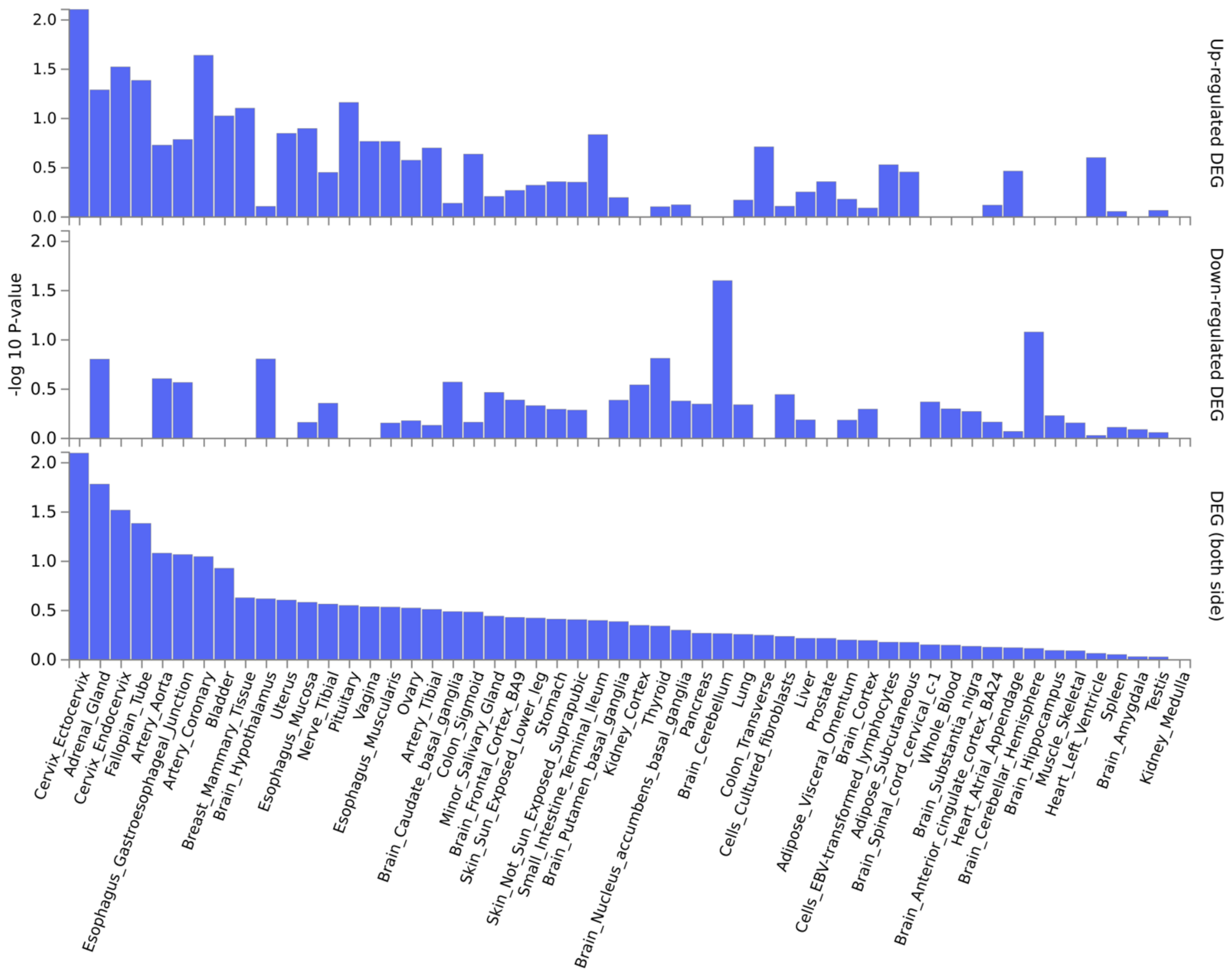

**Supplementary Figure 15: Tissue specificity analysis of enrichment of differentially expressed gene sets (DEG).**

Unadjusted p-values of enrichment of genes from SuSiE finemapping of GDM GWAS in gene sets for DEG in 54 tissue types in GTEx v8 using FUMA GENE2FUNC tool per Online Methods. Panels correspond to enrichment for genes with (A) higher expression in a given tissue, (B) lower expression in a given tissue, (C) any expression change in a given tissue. No results are significant after correction for multiple testing. Data available in Supplementary Table 13.

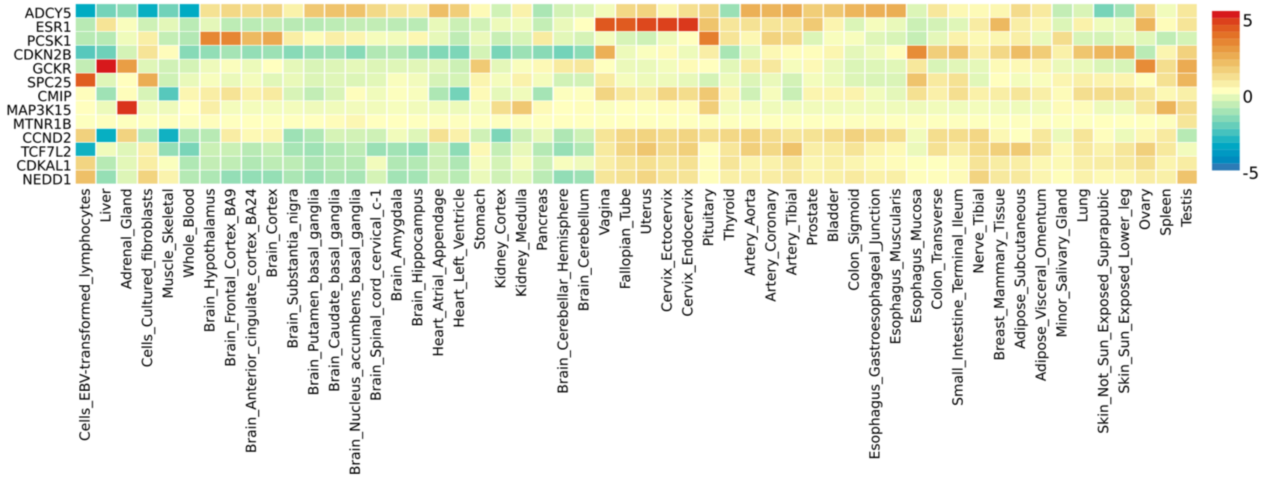

**Supplementary Figure 16: Heat map of gene expression.**
For each gene from finemapped GDM-associated loci, average log2(expression) in the given tissue compared to the average across tissues. Color scale is for intensity and direction of relative expression for gene labelled on Y-axis in tissue on X axis. Expression data from 54 tissues in GTEx analyzed using FUMA GENE2FUNC.

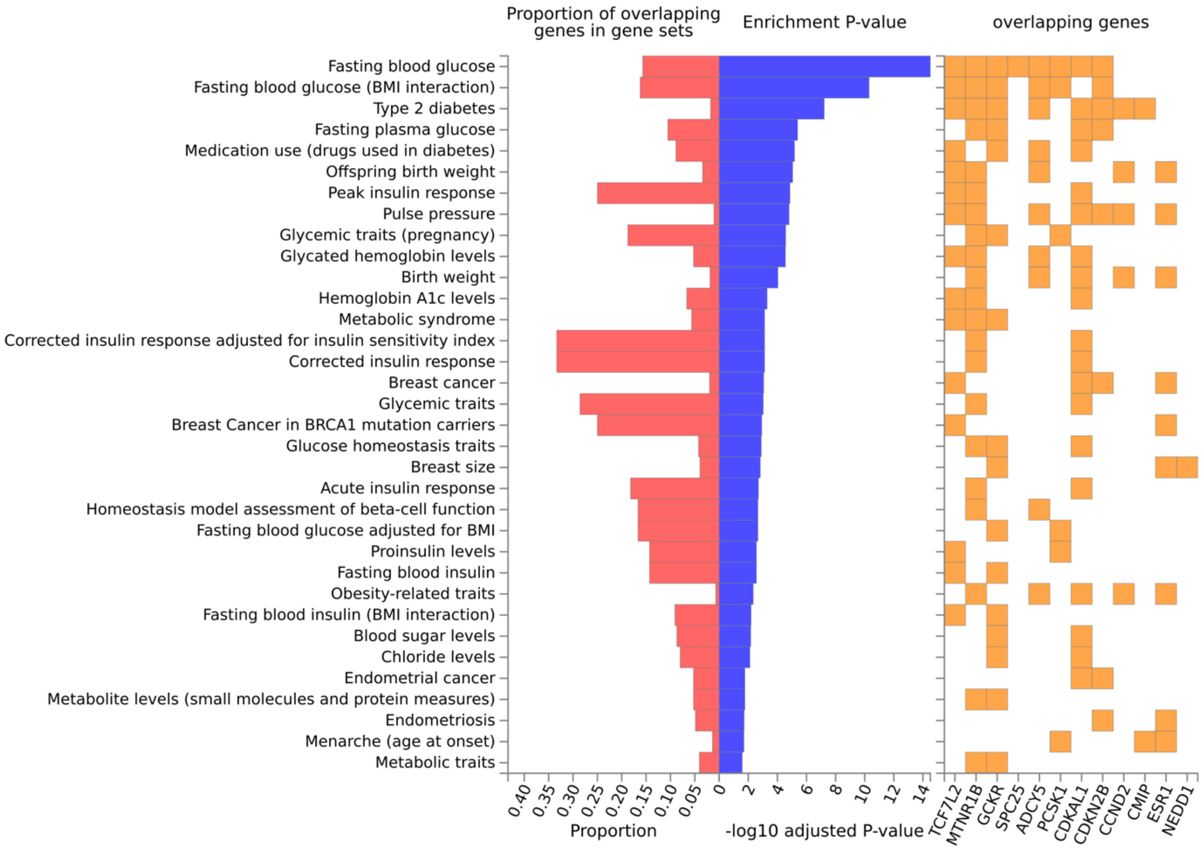

**Supplementary Figure 17: Enrichment of annotated genes for finemapped GDM loci in GWAS Catalog gene sets.**
Figures display gene sets with adjusted P-value < 0.05. Red bars indicate proportion of genes in the gene set that are among the GDM-related genes. Blue bars are the p-values after Bonferroni correction for number of tests. Yellow squares indicate overlapping genes. Performed using FUMA GENE2Func [https://fuma.ctglab.nl/tutorial#snp2gene]. Data available in Supplementary Table 14.

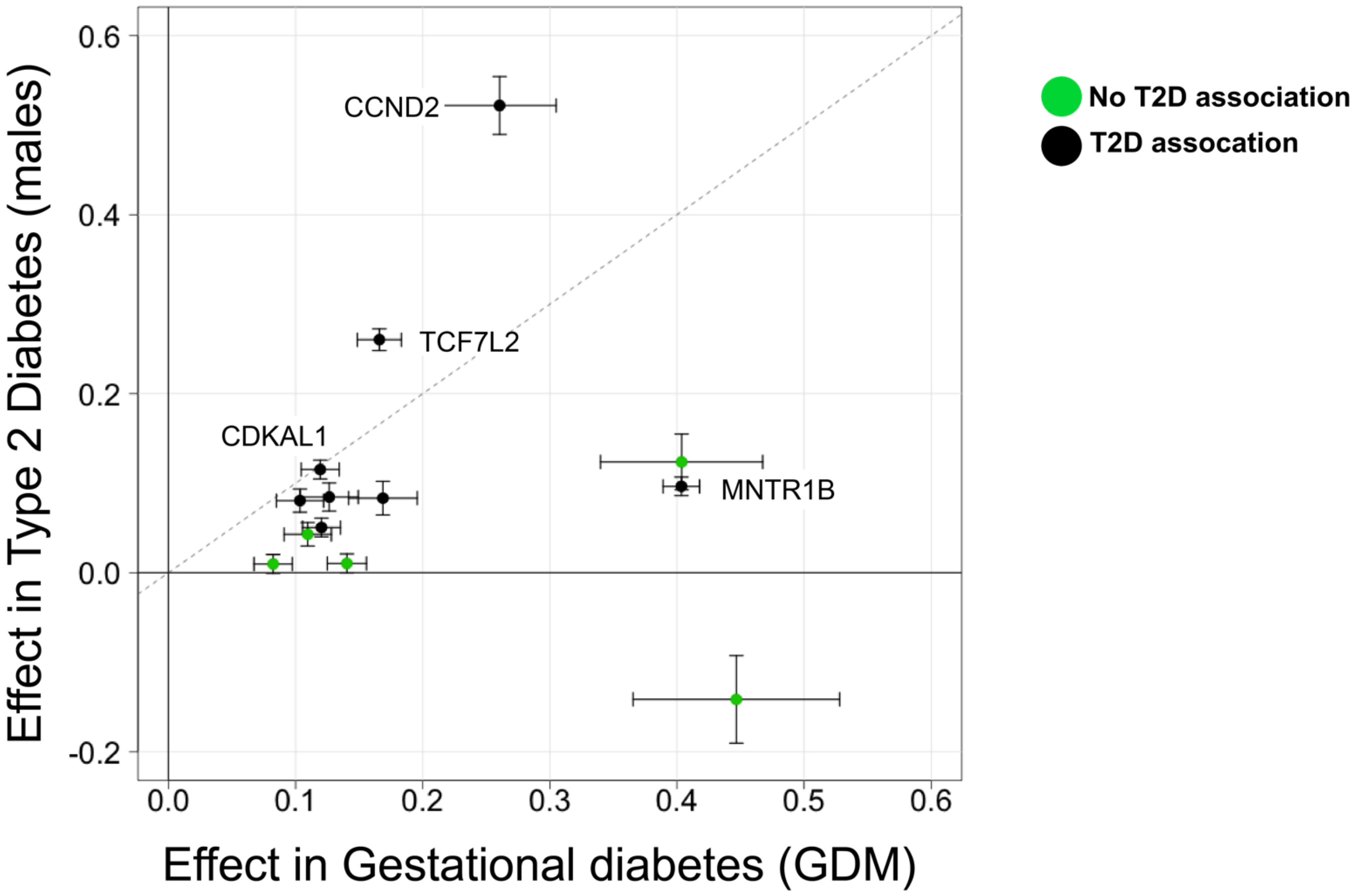

**Supplementary Figure 18: Genetic effect comparison in GDM vs T2D for GDM-associated loci**
Comparison of genetic effects in GWAS of GDM (x-axis) and T2D in males (y-axis). Heterogeneous appearance of effects was evident. The potential for components of effect was raised on gross examination – particularly given the more uniform appearance of many of the loci previously associated with T2D (green).

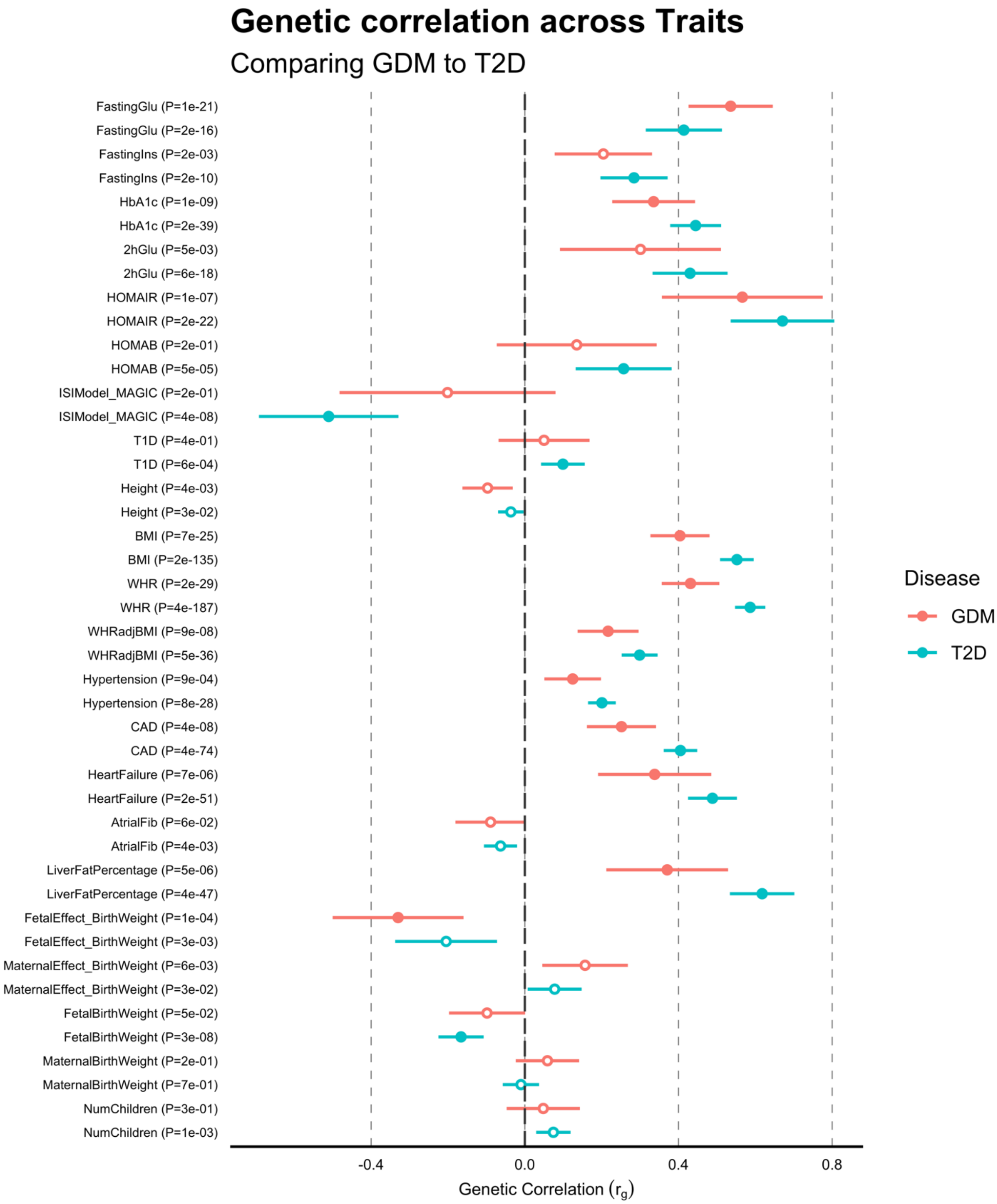

**Supplementary Figure 19: Genetic correlation of GDM with 22 Traits.**
Genetic correlations (SNP- $r_g$) of each trait with GDM (red) and T2D (Blue) estimated with LD score regression (Supplementary Table 14). Error bars reflect ±1 standard error. Filled points indicate the genetic correlation is significant after Bonferroni correction for multiple testing for both traits and biomarkers. The difference in $r_g$ between GDM and T2D is significant in 3 traits after Bonferroni correction: BMI, Waist-to-hip ratio adjusted for BMI, and CAD (Supplementary Table 15).

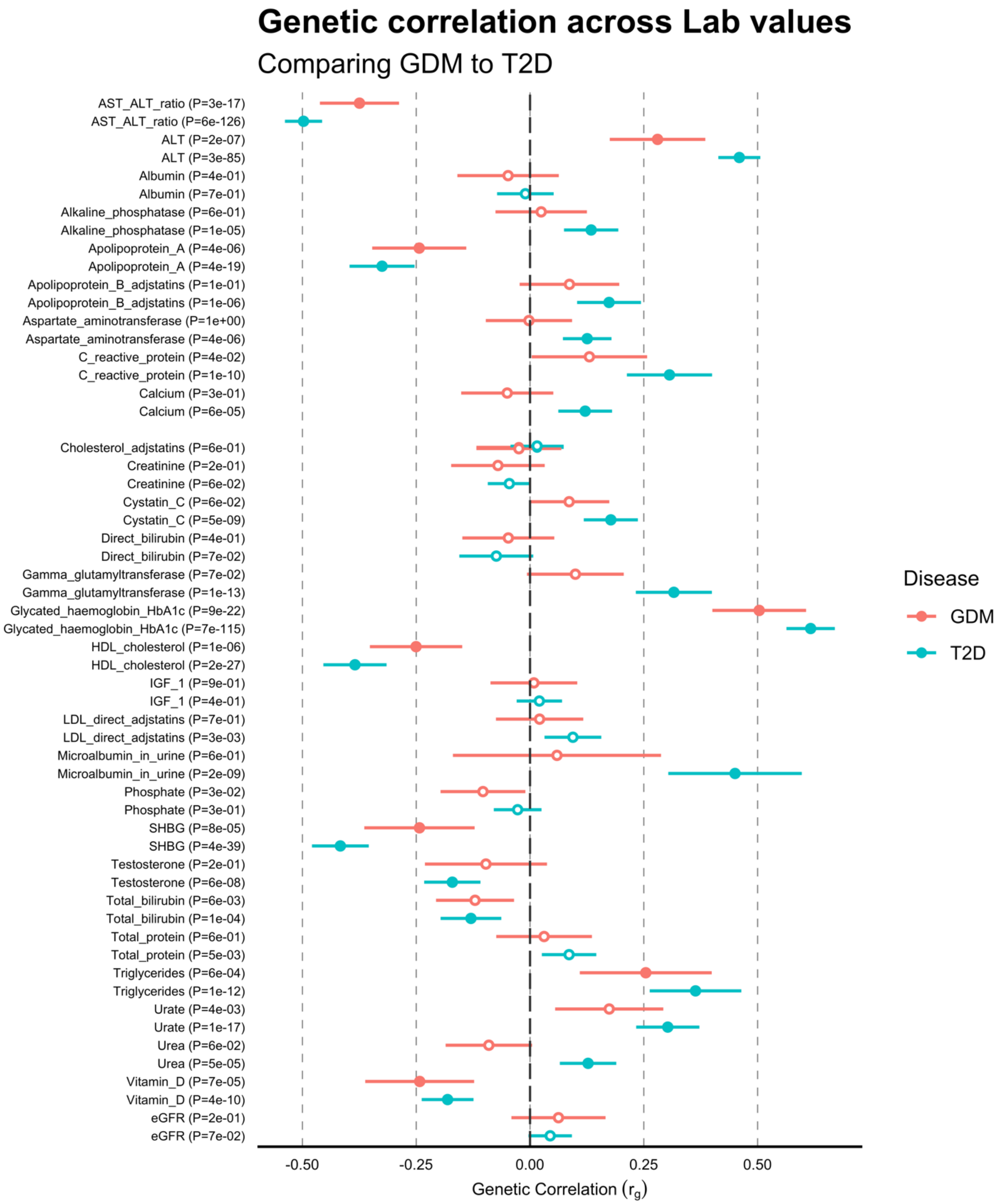

**Supplementary Figure 20: Genetic correlation of GDM with 29 biomarkers.**
Genetic correlations (SNP- $r_g$ ) of each biomarker with GDM (red) and T2D (Blue) estimated with LD score regression (Supplementary Table 15). Error bars reflect ±1 standard error. Filled points indicate the genetic correlation is significant after Bonferroni correction for multiple testing for both traits and biomarkers. The difference in $r_g$ between GDM and T2D is significant in 2 biomarkers after Bonferroni correction: gamma glutyl transferase and urea (Supplementary Table 16).

(A) Fasting glucose

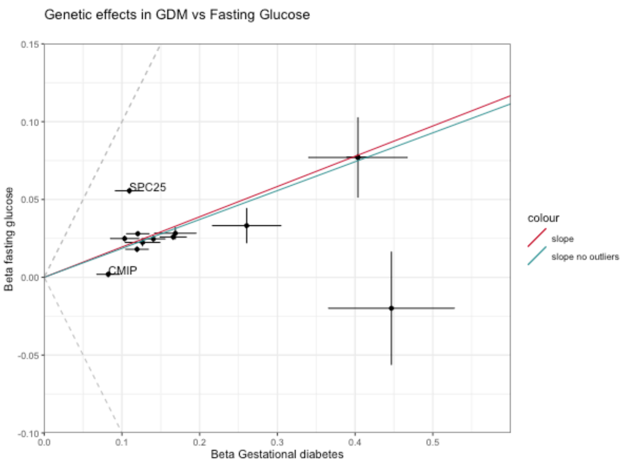

(B) 2h glucose

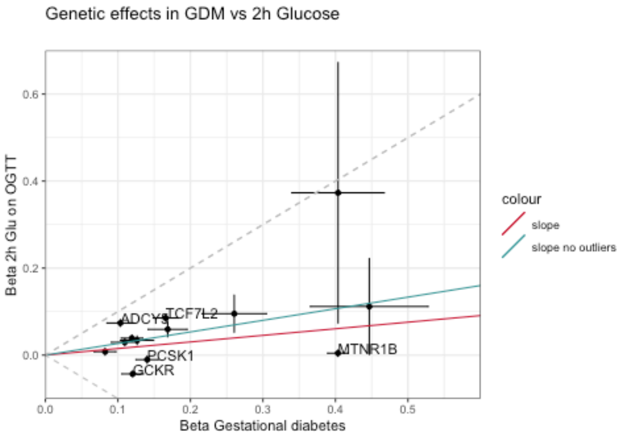

(C) HbA1C

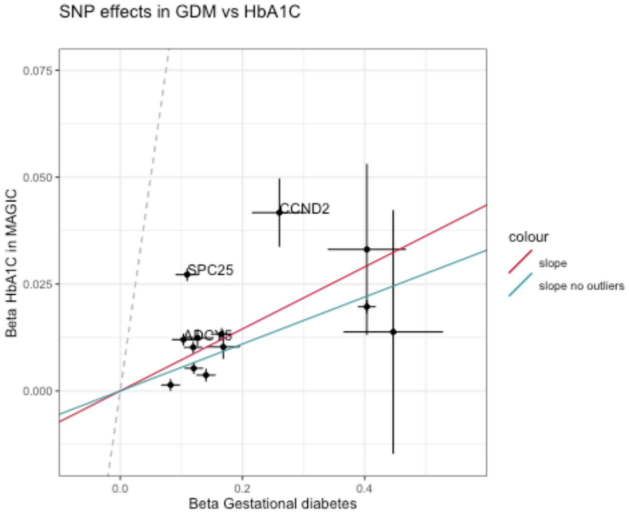

(D) Fasting insulin

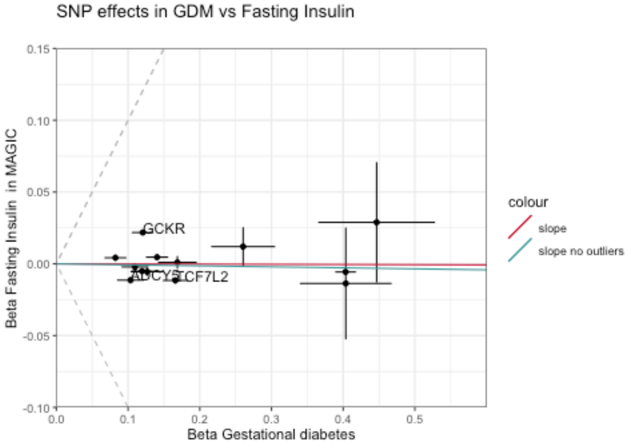

**Supplementary Figure 21: Comparison of effects of GDM-associated loci in GDM vs. glycemic traits**

Beta (log odds ratio) from the GWAS of GDM for each of the 13 significantly associated loci is compared to the beta from previous published GWAS of (A) fasting glucose, (B) 2 hour glucose, (C) HbA1C, and (D) fasting insulin. Error bars indicate +/- 1 standard error in each GWAS. Points labelled with gene names are identified as outliers by SCOUTJOY. Fitted slopes are shown from York regression with all variants (red) and after removal of identified outliers (blue). Dashed gray reference lines indicate equal absolute effect size. SCOUTJOY analysis suggests significant heterogeneity in all 4 comparisons, and a significant positive relationship of GDM with fasting glucose, 2 hour glucose, and HbA1C after outlier removal (Supplementary Table 20).

(A) Men

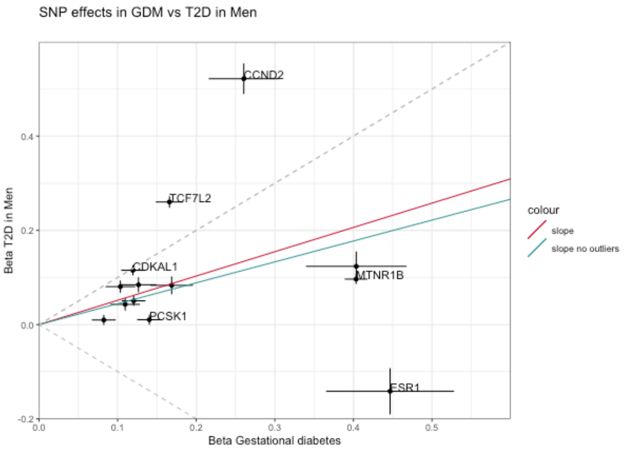

(B) Females

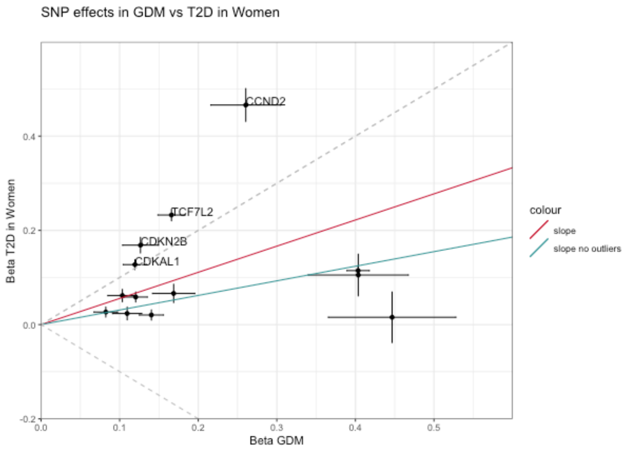

(C) Parous Females

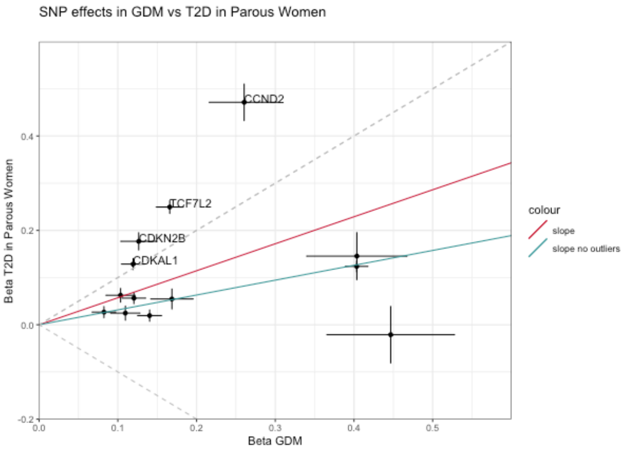

(D) Nulliparous Females

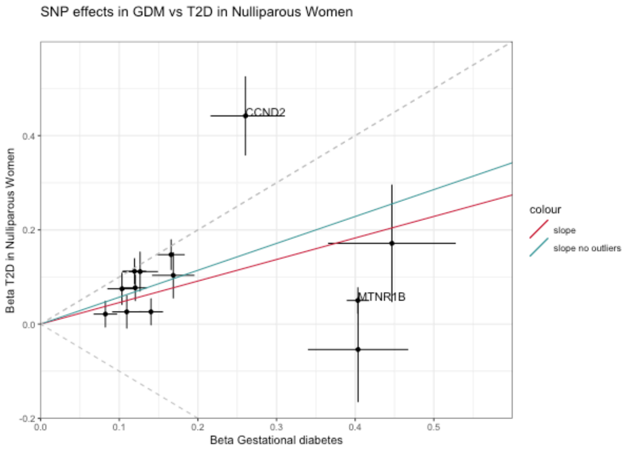

**Supplementary Figure 22: Comparison of effects of GDM-associated loci in GDM vs. T2D subgroups by sex and pregnancy history**

Betas (log odds ratios) from the GWAS of GDM for each of the 13 significantly associated loci are compared to the betas from GWAS of T2D in FinnGen among (A) males, (B) females, (C) parous females, and (D) nulliparous females. Error bars indicate +/- 1 standard error in each GWAS. Points labelled with gene names are identified as outliers by SCOUTJOY. Fitted slopes are shown from York regression with all variants (red) and after removal of identified outliers (blue). Dashed gray reference lines indicate equal absolute effect size. Data available in Supplementary Table 21

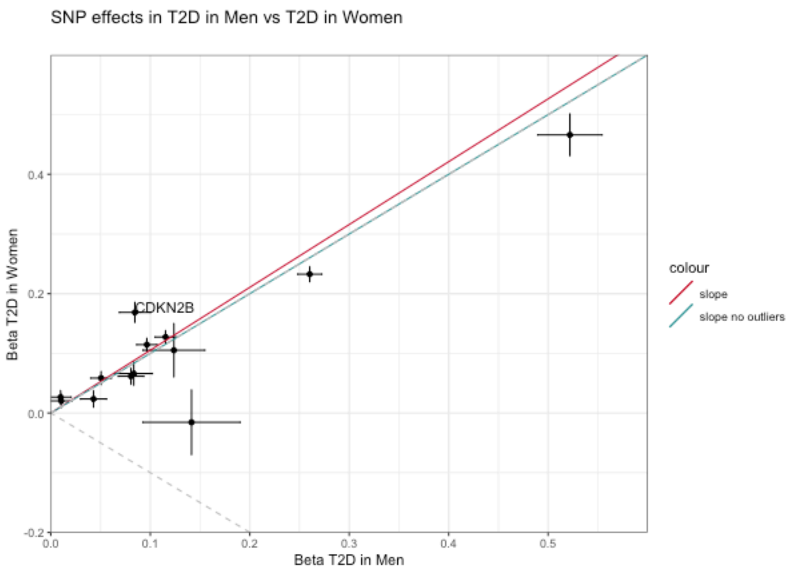

**Supplementary Figure 23: Comparison of effects of GDM-associated loci in T2D in males vs T2D in females**

Betas (log odds ratios) are reported from the GWAS of T2D in each sex in FinnGen. Error bars indicate +/- 1 standard error in each GWAS. Points labelled with gene names are identified as outliers by SCOUTJOY. Fitted slopes are shown from York regression with all variants (red) and after removal of identified outliers (blue). Dashed gray reference lines indicate equal absolute effect size. Data available in Supplementary Table 21

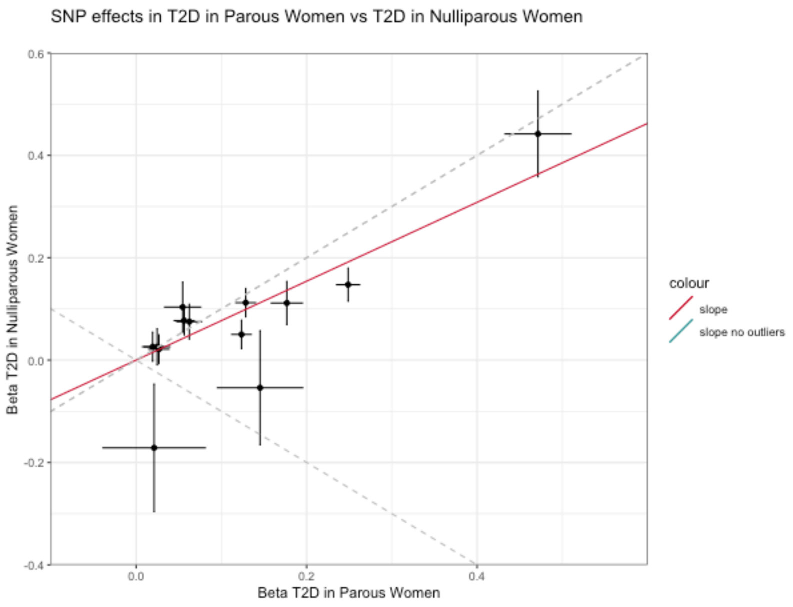

**Supplementary Figure 24: Comparison of effects of GDM-associated loci in T2D in parous females vs T2D in nulliparous females**

Betas (log odds ratios) are reported from the GWAS of T2D in parous and nulliparous females, respectively, in FinnGen. Error bars indicate +/- 1 standard error in each GWAS. No significant outliers are identified by SCOUTJOY. Fitted slope is from York regression with all variants. Dashed gray reference lines indicate equal absolute effect size. Data available in Supplementary Table 21

(A)

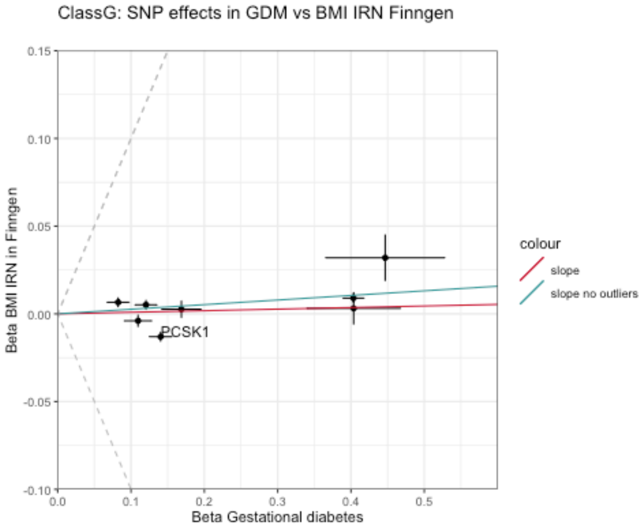

(B)

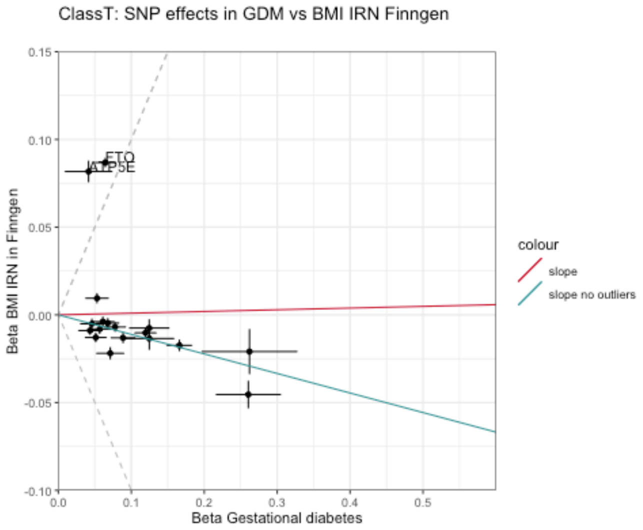

(C)

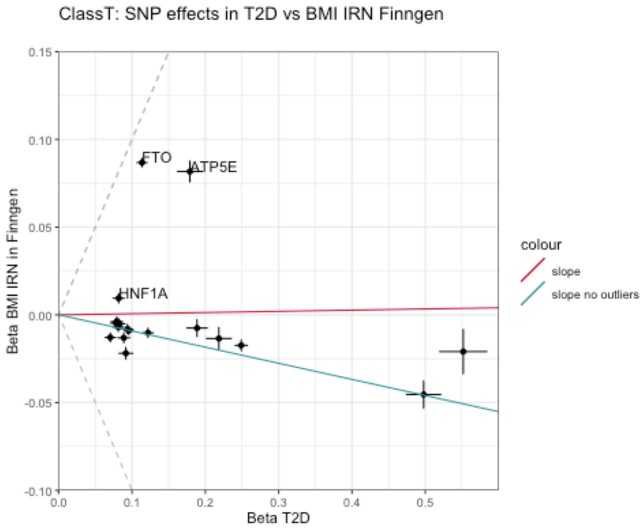

**Supplementary Figure 25: Comparison of GDM vs BMI effects sizes by GDM variant class**
Betas (log odds ratios) from the GWAS of GDM are compared to the betas from GWAS of inverse rank normalized (IRN) BMI in FinnGen among (A) ClassG SNPs and (B) ClassT SNPs from the shared variants analysis of GDM and T2D. For comparison, (C) IRN BMI effect sizes are also compared to GWAS of T2D in FinnGen. Error bars indicate +/- 1 standard error in each GWAS. Points labelled with gene names are identified as outliers by SCOUTJOY. Fitted slopes are shown from York regression with all variants (red) and after removal of identified outliers (blue). Dashed gray reference lines indicate equal absolute effect size. Data available in Supplementary Table 25.

(A) GDM

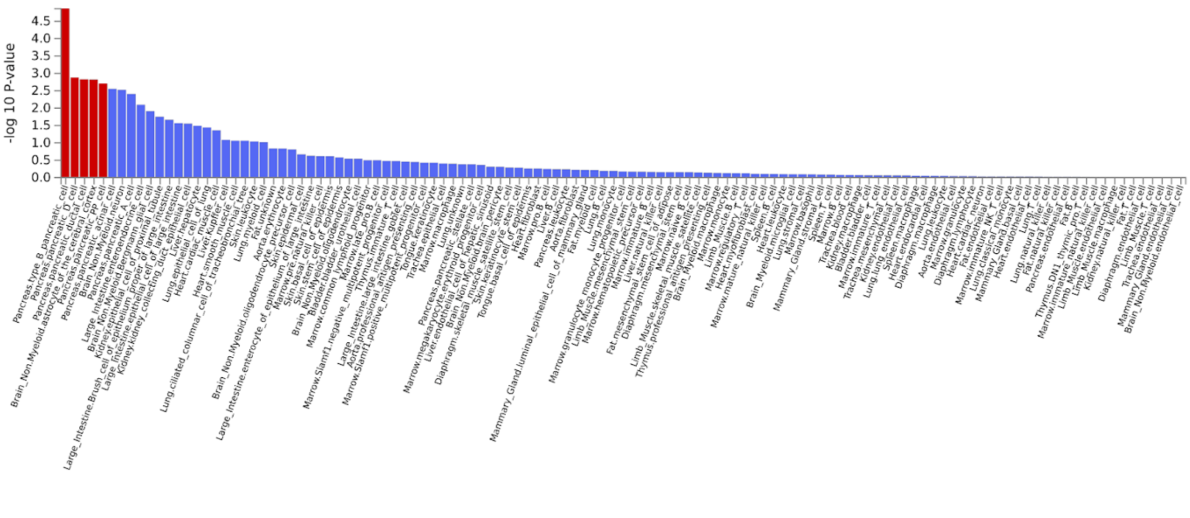

(B) T2D

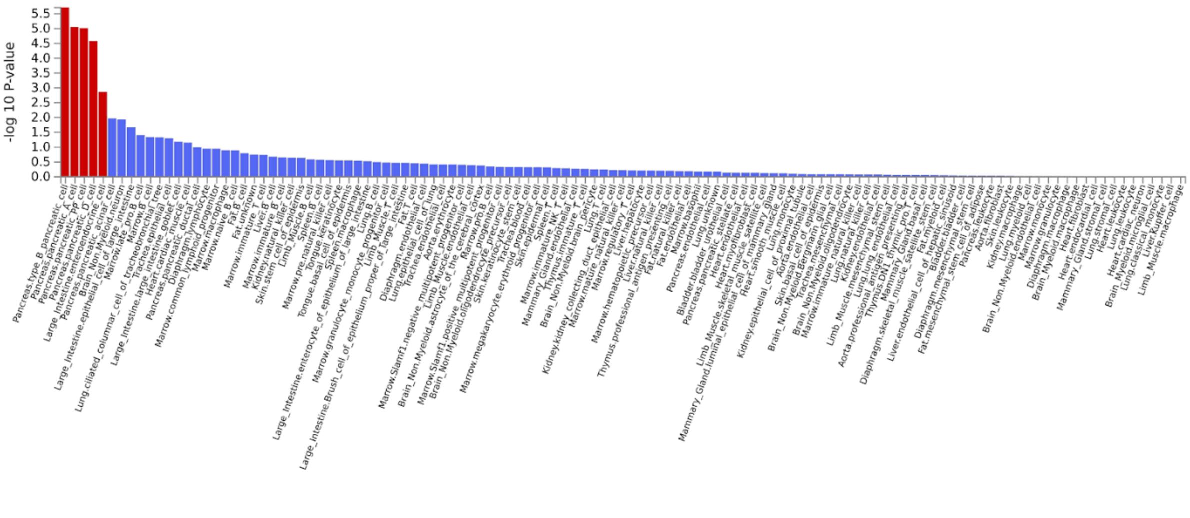

**Supplementary Figure 26: Cell type specificity analysis with GDM and T2D summary statistics**
Cell specificity analysis was performed for (A) the current GWAS of GDM and (B) published GWAS meta-analysis of T2D[26]. Unadjusted p-values are shown for the test of association between the GWAS results and results for cell-type specific gene expression for each cell-tissue pair. Analysis was performed using FUMA GENE2FUNC with murine single cell gene expression data from Tabula Muris. Cell-tissue pairs that remain significant after multiple testing correction are indicated in red. Data is available in Supplementary Table 26

(A) GDM

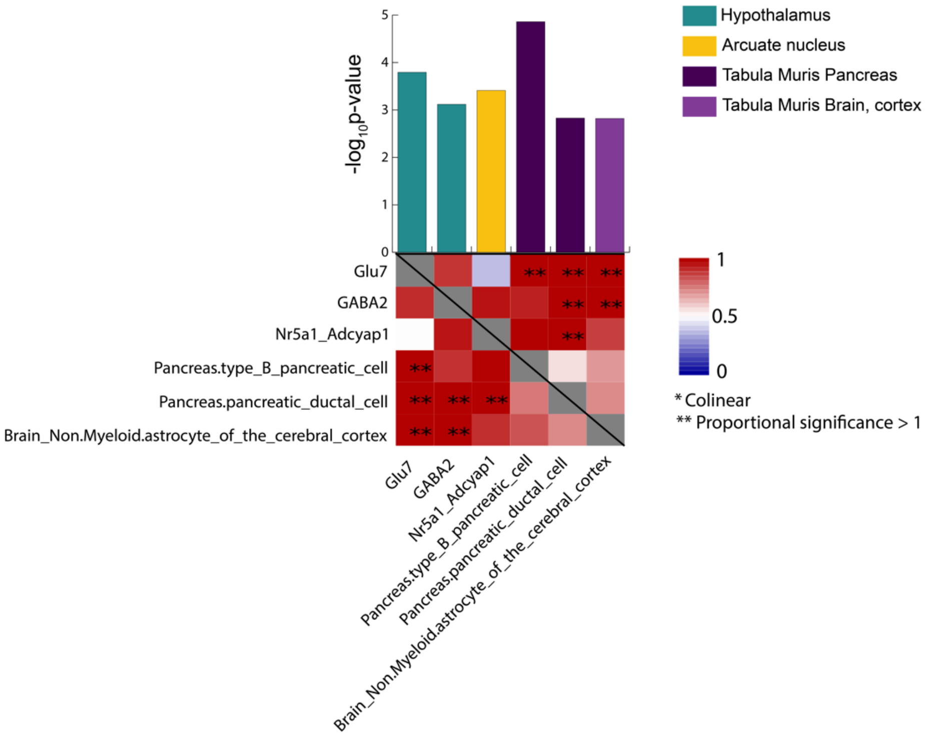

(B) T2D

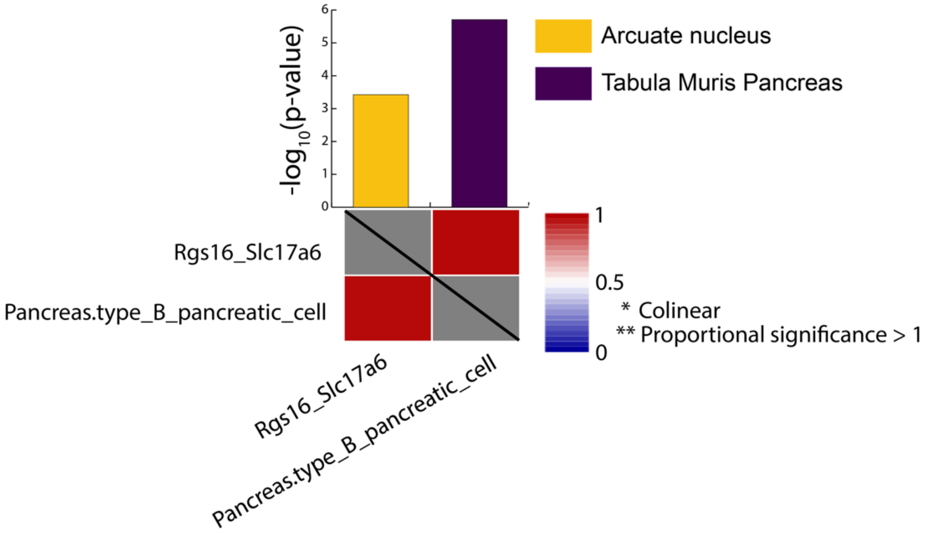

**Supplementary Figure 27: Cross dataset conditional analyses reveal independent signals for GDM and for T2D**

Evaluation of whether cell type specificity results across single cell expression datasets reflect overlapping genetic signals (Supplementary Note). Barplots indicate unadjusted P-values for the marginal association between relative gene expression in the given cell type and MAGMA gene-level associations in the GWAS of (A) GDM or (B) T2D. Colors indicate RNA-seq dataset source. Results are shown for cell types that are significantly associated with the GWAS after correction for multiple testing and have putatively independent association conditional on other cell types in the same RNA-seq dataset. Heatmaps are asymmetric and indicate the proportional significance (PS) of association with the cell type in the column after conditioning on the cell type in the row. Heatmap cell colors reflect PS between 0 and 1, with PS>1 is represented by double stars. Analysis was performed using FUMA GENE2FUNC. Data is available in Supplementary Table 28.

(A) Mouse pancreas: (B) Human pancreas:

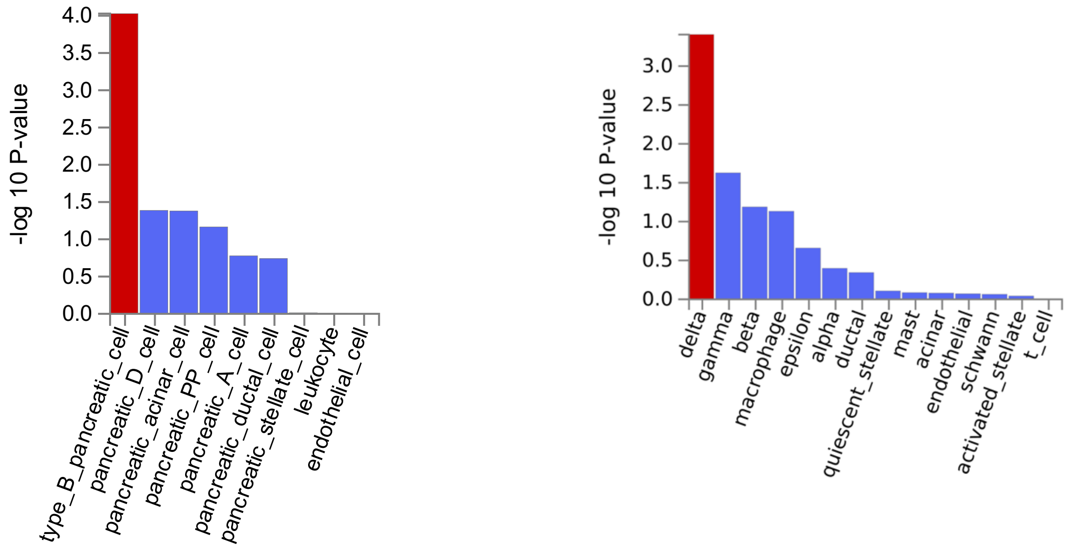

**Supplementary Figure 28: Cell type specificity analysis with GDM summary statistics in mouse vs human islets**

Cell specificity analysis for GDM GWAS with single cell gene expression data from (A) mouse pancreas in Tabula Muris and (B) a large single cell study of human pancreas[30]. Unadjusted p-values are shown for the test of association between the GWAS results and results for cell-type specific gene expression for each cell type. Cell types that remain significant after multiple testing correction are indicated in red. Analysis was performed using FUMA GENE2FUNC. Data is available in Supplementary Table 29.

Rayner, N.W., Qiao, Z., Moen, G.-H., Vaudel, M., Marsit, C.J., Chen, J., Nodzenski, M., Schnurr, T.M., Zafarmand, M.H., Bradfield, J.P., Grarup, N., Kooijman, M.N., Li-Gao, R., Geller, F., Ahluwalia, T.S., Paternoster, L., Rueedi, R., Huikari, V., Hottenga, J.-J., Lyytikäinen, L.-P., Cavadino, A., Metrustry, S., Cousminer, D.L., Wu, Y., Thiering, E., Wang, C.A., Have, C.T., Vilor-Tejedor, N., Joshi, P.K., Painter, J.N., Ntalla, I., Myhre, R., Pitkänen, N., van Leeuwen, E.M., Joro, R., Lagou, V., Richmond, R.C., Espinosa, A., Barton, S.J., Inskip, H.M., Holloway, J.W., Santa-Marina, L., Estivill, X., Ang, W., Marsh, J.A., Reichetzeder, C., Marullo, L., Hocher, B., Lunetta, K.L., Murabito, J.M., Relton, C.L., Kogevinas, M., Chatzi, L., Allard, C., Bouchard, L., Hivert, M.-F., Zhang, G., Muglia, L.J., Heikkinen, J., Consortium, E.G.G., Morgen, C.S., van Kampen, A.H.C., van Schaik, B.D.C., Mentch, F.D., Langenberg, C., Luan, J.a., Scott, R.A., Zhao, J.H., Hemani, G., Ring, S.M., Bennett, A.J., Gaulton, K.J., Fernandez-Tajes, J., van Zuydam, N.R., Medina-Gomez, C., de Haan, H.G., Rosendaal, F.R., Kutalik, Z., Marques-Vidal, P., Das, S., Willemsen, G., Mbarek, H., Müller-Nurasyid, M., Standl, M., Appel, E.V.R., Fonvig, C.E., Trier, C., van Beijsterveldt, C.E.M., Murcia, M., Bustamante, M., Bonas-Guarch, S., Hougaard, D.M., Mercader, J.M., Linneberg, A., Schraut, K.E., Lind, P.A., Medland, S.E., Shields, B.M., Knight, B.A., Chai, J.-F., Panoutsopoulou, K., Bartels, M., Sánchez, F., Stokholm, J., Torrents, D., Vinding, R.K., Willems, S.M., Atalay, M., Chawes, B.L., Kovacs, P., Prokopenko, I., Tuke, M.A., Yaghootkar, H., Ruth, K.S., Jones, S.E., Loh, P.-R., Murray, A., Weedon, M.N., Tönjes, A., Stumvoll, M., Michaelsen, K.F., Eloranta, A.-M., Lakka, T.A., van Duijn, C.M., Kiess, W., Körner, A., Niinikoski, H., Pahkala, K., Raitakari, O.T., Jacobsson, B., Zeggini, E., Dedoussis, G.V., Teo, Y.-Y., Saw, S.-M., Montgomery, G.W., Campbell, H., Wilson, J.F., Vrijkotte, T.G.M., Vrijheid, M., de Geus, E.J.C.N., Hayes, M.G., Kadarmideen, H.N., Holm, J.-C., Beilin, L.J., Pennell, C.E., Heinrich, J., Adair, L.S., Borja, J.B., Mohlke, K.L., Eriksson, J.G., Widén, E.E., Hattersley, A.T., Spector, T.D., Kähönen, M., Viikari, J.S., Lehtimäki, T., Boomsma, D.I., Sebert, S., Vollenweider, P., Sørensen, T.I.A., Bisgaard, H., Bønnelykke, K., Murray, J.C., Melbye, M., Nohr, E.A., Mook-Kanamori, D.O., Rivadeneira, F., Hofman, A., Felix, J.F., Jaddoe, V.W.V., Hansen, T., Pisinger, C., Vaag, A.A., Pedersen, O., Uitterlinden, A.G., Järvelin, M.-R., Power, C., Hyppönen, E., Scholtens, D.M., Lowe, W.L., Davey Smith, G., Timpson, N.J., Morris, A.P., Wareham, N.J., Hakonarson, H., Grant, S.F.A., Frayling, T.M., Lawlor, D.A., Njølstad, P.R., Johansson, S., Ong, K.K., McCarthy, M.I., Perry, J.R.B., Evans, D.M. & Freathy, R.M. Maternal and fetal genetic effects on birth weight and their relevance to cardio-metabolic risk factors. *Nature Genetics* **51**, 804-814 (2019).

17. York, D. Least squares fitting of a straight line with correlated errors. *Earth and Planetary Science Letters* **5**, 320-324 (1968).
18. York, D., Evensen, N.M., Martínez, M.L. & Delgado, J.D.B. Unified equations for the slope, intercept, and standard errors of the best straight line. *American Journal of Physics* **72**, 367-375 (2004).
19. Mahon, K.I. The New “York” Regression: Application of an Improved Statistical Method to Geochemistry. *International Geology Review* **38**, 293-303 (1996).
20. Verbanck, M., Chen, C.-Y., Neale, B. & Do, R. Detection of widespread horizontal pleiotropy in causal relationships inferred from Mendelian randomization between complex traits and diseases. *Nature Genetics* **50**, 693-698 (2018).
21. Titterington, D.M. & Halliday, A.N. On the fitting of parallel isochrons and the method of maximum likelihood. *Chemical Geology* **26**, 183-195 (1979).
22. Turley, P., Walters, R.K., Maghzian, O., Okbay, A., Lee, J.J., Fontana, M.A., Nguyen-Viet, T.A., Wedow, R., Zacher, M., Furlotte, N.A., Agee, M., Alipanahi, B., Auton, A., Bell, R.K., Bryc, K., Elson, S.L., Fontanillas, P., Furlotte, N.A., Hinds, D.A., Hromatka, B.S., Huber, K.E., Kleinman, A., Litterman, N.K., McIntyre, M.H., Mountain, J.L., Northover, C.A.M., Sathirapongsasuti, J.F., Sazonova, O.V., Shelton, J.F., Shringarpure, S., Tian, C., Tung, J.Y., Vacic, V., Wilson, C.H., Pitts, S.J., Magnusson, P., Oskarsson, S., Johannesson, M., Visscher, P.M., Laibson, D., Cesarini, D., Neale, B.M., Benjamin, D.J., andMe Research, T. & Social Science Genetic Association, C. Multi-trait analysis of genome-wide association summary statistics using MTAG. *Nature Genetics* **50**, 229-237 (2018).

23. North, B.V., Curtis, D. & Sham, P.C. A Note on the Calculation of Empirical P Values from Monte Carlo Procedures. *The American Journal of Human Genetics* **71**, 439-441 (2002).
24. Martin, J., Khramtsova, E.A., Goleva, S.B., Blokland, G.A.M., Traglia, M., Walters, R.K., Hubel, C., Coleman, J.R.I., Breen, G., Borglum, A.D., Demontis, D., Grove, J., Werge, T., Bralten, J., Bulik, C.M., Lee, P.H., Mathews, C.A., Peterson, R.E., Winham, S.J., Wray, N., Edenberg, H.J., Guo, W., Yao, Y., Neale, B.M., Faraone, S.V., Petryshen, T.L., Weiss, L.A., Duncan, L.E., Goldstein, J.M., Smoller, J.W., Stranger, B.E., Davis, L.K. & Sex Differences Cross-Disorder Analysis Group of the Psychiatric Genomics, C. Examining Sex-Differentiated Genetic Effects Across Neuropsychiatric and Behavioral Traits. *Biol Psychiatry* **89**, 1127-1137 (2021).
25. Karaderi, T., Drong, A.W. & Lindgren, C.M. Insights into the Genetic Susceptibility to Type 2 Diabetes from Genome-Wide Association Studies of Obesity-Related Traits. *Current Diabetes Reports* **15**, 83 (2015).
26. Mahajan, A., Taliun, D., Thurner, M., Robertson, N.R., Torres, J.M., Rayner, N.W., Payne, A.J., Steinthorsdottir, V., Scott, R.A., Grarup, N., Cook, J.P., Schmidt, E.M., Wuttke, M., Sarnowski, C., Magi, R., Nano, J., Gieger, C., Trompet, S., Lecoeur, C., Preuss, M.H., Prins, B.P., Guo, X., Bielak, L.F., Below, J.E., Bowden, D.W., Chambers, J.C., Kim, Y.J., Ng, M.C.Y., Petty, L.E., Sim, X., Zhang, W., Bennett, A.J., Bork-Jensen, J., Brummett, C.M., Canouil, M., Ec Kardt, K.U., Fischer, K., Kardia, S.L.R., Kronenberg, F., Lall, K., Liu, C.T., Locke, A.E., Luan, J., Ntalla, I., Nylander, V., Schonherr, S., Schurmann, C., Yengo, L., Bottinger, E.P., Brandslund, I., Christensen, C., Dedoussis, G., Florez, J.C., Ford, I., Franco, O.H., Frayling, T.M., Giedraitis, V., Hackinger, S., Hattersley, A.T., Herder, C., Ikram, M.A., Ingelsson, M., Jorgensen, M.E., Jorgensen, T., Kriebel, J., Kuusisto, J., Ligthart, S., Lindgren, C.M., Linneberg, A., Lyssenko, V., Mamakou, V., Meitinger, T., Mohlke, K.L., Morris, A.D., Nadkarni, G., Pankow, J.S., Peters, A., Sattar, N., Stancakova, A., Strauch, K., Taylor, K.D., Thorand, B., Thorleifsson, G., Thorsteinsdottir, U., Tuomilehto, J., Witte, D.R., Dupuis, J., Peyser, P.A., Zeggini, E., Loos, R.J.F., Froguel, P., Ingelsson, E., Lind, L., Groop, L., Laakso, M., Collins, F.S., Jukema, J.W., Palmer, C.N.A., Grallert, H., Metspalu, A., Dehghan, A., Kottgen, A., Abecasis, G.R., Meigs, J.B., Rotter, J.I., Marchini, J., Pedersen, O., Hansen, T., Langenberg, C., Wareham, N.J., Stefansson, K., Gloyn, A.L., Morris, A.P., Boehnke, M. & McCarthy, M.I. Fine-mapping type 2 diabetes loci to single-variant resolution using high-density imputation and islet-specific epigenome maps. *Nat Genet* **50**, 1505-1513 (2018).
27. Tabula Muris Consortium. Single-cell transcriptomics of 20 mouse organs creates a Tabula Muris. *Nature* **562**, 367-372 (2018).
28. Chen, R., Wu, X., Jiang, L. & Zhang, Y. Single-Cell RNA-Seq Reveals Hypothalamic Cell Diversity. *Cell Rep* **18**, 3227-3241 (2017).
29. Campbell, J.N., Macosko, E.Z., Fenselau, H., Pers, T.H., Lyubetskaya, A., Tenen, D., Goldman, M., Verstegen, A.M., Resch, J.M., McCarroll, S.A., Rosen, E.D., Lowell, B.B. & Tsai, L.T. A molecular census of arcuate hypothalamus and median eminence cell types. *Nat Neurosci* **20**, 484-496 (2017).
30. Baron, M., Veres, A., Wolock, S.L., Faust, A.L., Gaujoux, R., Vetere, A., Ryu, J.H., Wagner, B.K., Shen-Orr, S.S., Klein, A.M., Melton, D.A. & Yanai, I. A Single-Cell Transcriptomic Map of the Human and Mouse Pancreas Reveals Inter- and Intra-cell Population Structure. *Cell Syst* **3**, 346-360 e4 (2016).
31. Watanabe, K., Umicevic Mirkov, M., de Leeuw, C.A., van den Heuvel, M.P. & Posthuma, D. Genetic mapping of cell type specificity for complex traits. *Nat Commun* **10**, 3222 (2019).
32. Ladyman, S.R. & Grattan, D.R. Region-Specific Suppression of Hypothalamic Responses to Insulin To Adapt to Elevated Maternal Insulin Secretion During Pregnancy. *Endocrinology* **158**, 4257-4269 (2017).
33. He, Y., Xu, P., Wang, C., Xia, Y., Yu, M., Yang, Y., Yu, K., Cai, X., Qu, N., Saito, K., Wang, J., Hyseni, I., Robertson, M., Piyarathna, B., Gao, M., Khan, S.A., Liu, F., Chen, R., Coarfa, C., Zhao, Z., Tong, Q., Sun, Z. & Xu, Y. Estrogen receptor-alpha expressing neurons in the ventrolateral VMH regulate glucose balance. *Nat Commun* **11**, 2165 (2020).

34. Yang, J.A., Stires, H., Belden, W.J. & Roepke, T.A. The Arcuate Estrogen-Regulated Transcriptome: Estrogen Response Element-Dependent and -Independent Signaling of ERalpha in Female Mice. *Endocrinology* **158**, 612-626 (2017).
35. Dirice, E., De Jesus, D.F., Kahraman, S., Basile, G., Ng, R.W., El Ouaamari, A., Teo, A.K.K., Bhatt, S., Hu, J. & Kulkarni, R.N. Human duct cells contribute to beta cell compensation in insulin resistance. *JCI Insight* **4**(2019).
36. Dirice, E., Basile, G., Kahraman, S., Diegisser, D., Hu, J. & Kulkarni, R.N. Single-nucleus RNA-Seq reveals singular gene signatures of human ductal cells during adaptation to insulin resistance. *JCI Insight* **7**(2022).
